# Symptom propagation in respiratory pathogens of public health concern: a review of the evidence

**DOI:** 10.1101/2024.01.05.24300898

**Authors:** Phoebe Asplin, Rebecca Mancy, Thomas Finnie, Fergus Cumming, Matt J. Keeling, Edward M. Hill

## Abstract

Symptom propagation occurs when the symptom set an individual experiences is correlated with the symptom set of the individual who infected them. Symptom propagation may dramatically affect epidemiological outcomes, potentially causing clusters of severe disease. Conversely, it could result in chains of mild infection, generating widespread immunity with minimal cost to public health.

Despite accumulating evidence that symptom propagation occurs for many respiratory pathogens, the underlying mechanisms are not well understood. Here we conducted a scoping literature review for 14 respiratory pathogens to ascertain the extent of evidence for symptom propagation by two mechanisms: dose-severity relationships and route-severity relationships.

We identify considerable heterogeneity between pathogens in the relative importance of the two mechanisms, highlighting the importance of pathogen-specific investigations. For almost all pathogens, including influenza and SARS-CoV-2, we found support for at least one of the two mechanisms. For some pathogens, including influenza, we found convincing evidence that both mechanisms contribute to symptom propagation.

Furthermore, infectious disease models traditionally do not include symptom propagation. We summarise the present state of modelling advancements to address the methodological gap. We then investigate a simplified disease outbreak scenario, finding that under strong symptom propagation, quarantining mildly infected individuals can have negative epidemiological implications.

## 1 Introduction

Respiratory pathogens can inflict a considerable burden on public health; in 2018, before the COVID-19 pandemic, 6.1% of total deaths in the UK were attributable to respiratory infections [1]. Though capable of causing substantial mortality, infection by respiratory pathogens can often result in differing severity amongst the population. The factors underlying symptom severity are yet to be fully understood. One facet of symptom severity is the concept of symptom propagation. In broad terms, symptom propagation is when the symptom set an individual experiences depends on the symptom set of the individual who infected them. One exemplar pathogen is *Yersinia pestis*, the bacterial causative agent of plague. It is well-documented that the symptoms experienced by an individual depend on the route of transmission via which they acquired infection, and thus on the severity of the individual who infected them. Specifically, for those who develop the most acute form, pneumonic plague, their onward air-borne infections lead to secondary cases also developing pneumonic plague [2]; conversely, those with bubonic plague are unable to transmit disease through the air-borne route [2]. There is growing evidence that a similar relationship may exist for other well-studied pathogens, such as SARS-CoV-2, either through a relationship between aerosol transmission and severity [3] or due to the relationship between infectious dose and severity [4]. The COVID-19 pandemic has resulted in a relative glut of literature relevant to symptom propagation [5]. These studies have led to a greater understanding of the underlying mechanisms of respiratory pathogens, allowing for questions surrounding symptom propagation to be answered with greater certainty and motivating more generic consideration of symptom propagation as an important epidemiological concept.

We regard symptom propagation to be of notable importance to public health. It is conceivable for symptom propagation to result in extreme epidemiological outcomes. On the one hand, large-scale clusters of severe infections may be generated that would be detrimental to those most at risk. On the other hand, propagation of symptom severity could lead to chains of mild or asymptomatic infections that generate widespread immunity with minimal cost to public health. There are also circumstances where symptom propagation may be harnessed to amplify the impact of public health measures, resulting in a reduction in symptom severity throughout the population and a lessened burden on public health resources.

Despite these potential impacts, symptom propagation is biologically and mathematically understudied. At the time of writing, prior research in related specialised research areas has been restricted to reviews on the aerosol transmission route [3, 6], dose-response relationships [4] and the effect of non-pharmaceutical interventions reducing disease severity [7]. The number of pathogens typically examined in these previous studies has also been limited, primarily focusing on influenza and most recently SARS-CoV-2 (as a consequence of the COVID-19 pandemic), with other pathogens of public health concern largely overlooked.

Detailed quantitative studies can determine the contexts where symptom propagation can have an amplifying or mitigating role in disease outbreaks. Compartmental models are a benchmark modelling paradigm in the mathematical modelling of infectious diseases. Based on a “standard” SIR model, with states representing susceptible, infectious and recovered disease states, there are extensions to incorporate additional structures that are commonplace (e.g. latent states, age, spatial variation) [8]. Symptom propagation is an understudied model extension; hence there is limited understand of when the propagation of symptom severity can lead to different courses of public health action being advised.

Through this review, we consolidate and synthesise evidence for the propagation of symptom severity for a broad range of respiratory pathogens of public health concern. We additionally consider how symptom propagation has previously been considered within mathematical models, and supplement this with a case study to show the importance of symptom propagation on evaluations of intervention strategies.

Our review is structured as follows. Section 2 contains preliminaries and definitions. Section 3 outlines our scoping review methodology. Section 4 describes our appraisal of the evidence for the propagation of symptom severity. We detail four pathogens in the main manuscript, influenza, SARS-CoV-2, measles and *Yersinia pestis*, with findings for ten more pathogens reported in the Supporting Information. Within Section 5, we then delve into epidemiological modelling and model frameworks that explicitly incorporate symptom propagation and present a case study where particular targeting of public health interventions can result in worsened epidemiological outcomes. To conclude, Section 6 summarises the scientific contribution of our review.

## 2 Background

Here we provide the basic definitions that will be used throughout this review, such that there is a consistent and robust definition of each term. We consider ‘symptom propagation’ (Section 2.1), ‘severe disease’ (Section 2.2) and the mechanisms of symptom propagation (Section 2.3), before providing a general glossary of other common terms (Section 2.4).

### 2.1 Defining symptom propagation

There are many factors that influence an individual’s symptom set, including the dose with which they are infected and the route of transmission through which this occurs. In turn, these factors are affected by other variables, such as the setting in which infection occurs (household versus community, poorly ventilated spaces, etc.), weather (including humidity, in particular), and the pathogen strain. Here, we review evidence that the symptom set of the infecting individual also affects the dose and transmission route, and therefore the symptom set experienced.

We define symptom propagation to be when the symptom set of an infected individual depends on the symptom set of the individual from which they acquired infection. We only consider symptom propagation to occur through epidemiological mechanisms and explicitly exclude pathogen heterogeneity through evolution. Although this idea can be applied to symptom sets in general, we have chosen to look specifically at symptom severity. Our reasoning for this choice is that whilst symptoms vary, all pathogens result in more and less severe forms of disease, and thus, symptom severity is more generally applicable. Symptom severity is also more directly relevant to public health and policy decisions.

### 2.2 Defining ‘severe’ disease

The literature does not currently provide a consistent scheme for defining disease severity, and variations are seen across studies investigating the same pathogen. There is even greater heterogeneity across pathogens, often motivated by their specific clinical presentations. For example, within infectious disease modelling for COVID-19, infected individuals are typically classed as either asymptomatic or symptomatic, where symptomatic includes both those with minimal symptoms that we would usually class as ‘mild’ (e.g. sore throat or runny nose) and those with severe, or even fatal, symptoms [9]. In contrast, in the case of plague, individuals are typically classed as having either bubonic or pneumonic plague, where both categorisations are associated with high mortality (although pneumonic plague is notably more fatal) [2]. In many cases, severity is a continuum, with the separation between mild and severe somewhat arbitrary.

Here, we have generally chosen to identify ‘mild’ disease with upper respiratory tract (URT) symptoms such as sore throat or a blocked/runny nose. Such symptoms are generally associated with the ‘common cold’ [10]. In contrast, we consider ‘severe’ disease to be associated with lower respiratory tract (LRT) symptoms such as difficulty breathing or severe cough. Whilst these symptoms align with our personal view of how to categorise ‘severe’ disease, we also chose this classification as LRT infection is often used as a marker for severe disease within hospitals [11, 12].

Furthermore, we acknowledge that symptom severity is a spectrum and symptom propagation is a set of mechanisms that determine correlations between the position on the severity spectrum of an infected individual with the individual from whom they contracted disease. Our contention, therefore, is that we would expect to see associations at all levels of symptom severity, from comparing asymptomatic to symptomatic individuals to comparing fatal and non-fatal hospitalised cases. Consequently, we still discuss studies with a large range of severity classifications, including more systemic markers of severe disease, such as fever.

### 2.3 Symptom propagation mechanisms

We distinguish between two mechanisms through which symptom propagation can occur (Fig. 1): dose-severity and route-severity relationships. We consider these two mechanisms independently throughout this review for simplicity. We consider these two mechanisms independently throughout this review for simplicity. However, we recognise that dose and route may not be independent of one other; for example, aerosol transmission typically results in a smaller infectious dose than close contact transmission [13]. This relationship is further complicated by the variation in the dose required for infection at different sites; for example, a lower dose is typically required to initiate infection in the LRT compared to the URT [13–15]. Ultimately, we would like to be able to compute the ‘effective dose’ associated with a transmission event (conditioning on the route of infection and site of infection). In the absence of clear information on this in the literature, we treat the dose-severity and route-severity relationships separately, acknowledging that this is a simplification.

**Fig. 1.**
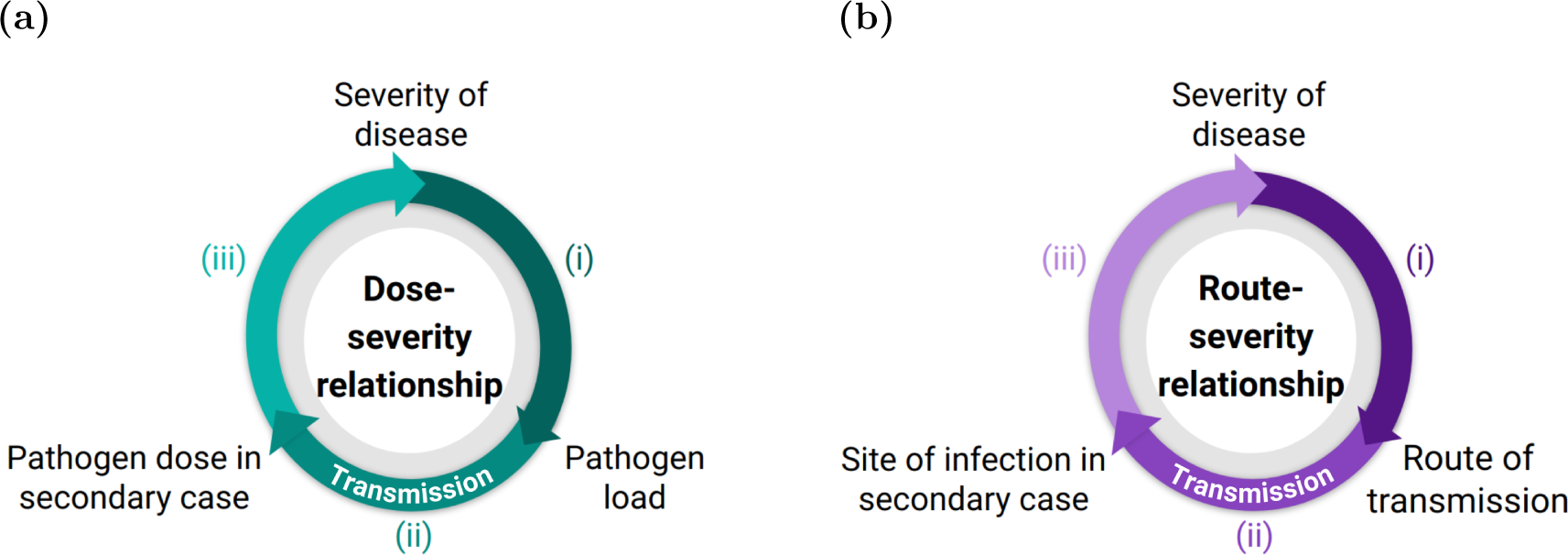
Cycle diagrams depicting two symptom propagation mechanisms: dose-severity relationships and route-severity relationships. (a) A dose-severity relationship arises when (i) an individual’s disease severity determines their pathogen load; (ii) pathogen load affects the infectious dose with which they infect others; (iii) this infectious dose then determines the disease severity in the secondary case. (b) A route-severity relationship arises when (i) an individual’s disease severity determines the transmission route through which they infect others; (ii) the transmission route then determines the site of infection in the secondary case; (iii) the site of infection then affects their disease severity.

#### Dose-severity relationships

A dose-severity relationship exists when pathogen load and symptom severity are positively correlated. Individuals with more severe disease tend to have a higher pathogen load [16]. A higher pathogen load leads to more pathogen being excreted such that those they infect tend to receive a larger infectious dose, increasing the probability of more severe disease outcomes [4].

In assessing whether a dose-severity mechanism is apparent for specific pathogens, measuring the pathogen load is an important step. Pathogen load is often measured directly through plaque assay or quantitative PCR (qPCR) [17, 18]. Another established way of capturing pathogen load is through the Cycle threshold (Ct) value, based on real-time PCR (polymerase chain reaction) assays [19]. Ct levels are inversely proportional to the amount of target nucleic acid in the sample and hence pathogen load. Therefore, we collectively considered studies looking either directly at pathogen load or at Ct values.

#### Route-severity relationships

Respiratory pathogens can generally be transmitted through multiple transmission routes. The number of categories of transmission route and their associated terms vary in the literature; however, the following three forms are most common [6, 20, 21]: (i) Aerosol transmission - infection via the inhalation of small droplets (*<* 5*µm*); (ii) Large droplet transmission - infection via the inhalation of large droplets (*≥* 5*µm*); (iii) Direct contact transmission - infection via contact with an infected individual or object (fomite).

Given we are predominantly interested in transmission routes with the potential for pathogens to enter the LRT, we often group the above three transmission routes into aerosol transmission (capable of penetrating the LRT) and close contact transmission (large droplet and direct contact, only capable of infecting the URT).

Those with severe disease (LRT infection) often produce a larger volume of aerosols and/or have increased capability to produce aerosols compared to those with mild disease (URT infection), who are instead more likely to transmit disease via close contact transmission [22–24]. Infection via aerosols is then associated with more severe symptoms, due to aerosols having the ability to reach the LRT [25, 26].

### 2.4 Glossary

#### Aerosols

Small droplets (*<* 5*µm*) that can be inhaled and reach the lower respiratory tract.

#### Close contact transmission

Transmission that requires individuals to be within a short distance of each other (i.e. large droplet or direct contact transmission).

#### Direct contact transmission

Infection direct contact with particles that are picked up onto an individual’s skin after direct contact with an infected individual or surface (fomite).

#### Case fatality rate (CFR)

The proportion of reported cases that are fatal.

#### Cycle threshold (Ct) value

For real-time PCR (polymerase chain reaction) assays, a positive reaction is detected by accumulation of a fluorescent signal. The Ct value is defined as the number of cycles required for the fluorescent signal to cross the threshold (i.e. exceeds background level). Ct levels are inversely proportional to the amount of target nucleic acid in the sample (the lower the Ct level, the greater the amount of target nucleic acid in the sample). Due to this inverse relationship, low Ct value corresponds to a high pathogen load and can be used as a proxy [27–29].

#### Exhaled breath condensate (EBC)

Cooled and condensed exhaled air, providing a non-invasive method of sampling airway lining fluid.

#### Infectious dose

The number of pathogens an individual is infected with, typically measured by colony forming units (CFU) and often stated with respect to the 50% tissue culture infectious dose (TCID_50_, the dilution of pathogen required to infect 50% of a cell culture).

#### Intranasal inoculation

Intentional infection of a human volunteer or animal via the nasal cavity, usually either through drops or sprays.

#### Large droplet

Droplets that are *≥* 5*µm* which can be inhaled but are generally too large to reach the LRT.

#### Lower respiratory tract (LRT)

Consists of the larynx, trachea, bronchi and the lungs.

#### Respiratory pathogen

Pathogens which can initiate infection in the respiratory tract.

#### Symptom propagation

When the symptom set of an infected individual depends on the symptom set of the individual from which they acquired infection. We only consider symptom propagation to occur through epidemiological mechanisms and explicitly exclude pathogen heterogeneity through evolution.

#### Upper respiratory tract (URT)

Consists of the nose, nasal cavity and the pharynx.

## 3 Search methodology

We performed a scoping literature review using Google Scholar and PubMed published up until August 2023. We included all peer-reviewed studies that were deemed relevant to symptom propagation; we did not consider pre-prints. Within the peer-reviewed literature we did not exclude any study types. We considered a total of 14 pathogens (listed alphabetically): Adenovirus, *Bordetella pertussis*, Group A streptococci, influenza, measles, MERS-CoV, *Mycobacterium tuberculosis*, RSV, Rhinovirus, SARS-CoV-1, SARS-CoV-2, Variola virus (Smallpox), Varicella zoster virus (chickenpox), *Yersinia pestis*. For each pathogen, we performed a number of separate searches focused on specific parts of the symptom propagation mechanisms.

The aim of this review is to demonstrate that there exists evidence for symptom propagation across a range of pathogens, and hence it is not an exhaustive review of the literature. The 14 pathogens in this study were chosen as pathogens capable of initiating infection in the respiratory tract (which we refer to as respiratory pathogens), being of public health concern (either presently or historically), and with the requirement of there being sufficient relevant studies in the literature.

For dose-response relationships, we performed three separate literature searches, each corresponding to part of the mechanism shown in Fig. 1. Our search terms were:

(i) (*pathogen name* OR *disease name*) AND (“viral” OR “bacterial”) AND (“load” OR “shedding”) AND (“severity” OR “symptoms”)
(ii) (*pathogen name* OR *disease name*) AND (“viral” OR “bacterial”) AND “load” AND (“shedding” OR “aerosol production”)
(iii) (*pathogen name* OR *disease name*) AND (“inoculant dose” OR “intensity of exposure”) AND (“severity” OR “symptoms”)

For route-severity relationships, we initially performed a search to determine the transmission routes and possible sites of initial infection for the pathogen (i.e. whether it could initiate infection in both the URT and LRT). Given evidence for both aerosol transmission and transmission via at least one other route, and evidence for initial infection occurring in both the URT and LRT, we searched for evidence of route-severity relationships. We performed two separate literature searches: evidence that severe disease leads to increased aerosol production ((i) in Fig. 1) and evidence that increased aerosol production leads to more severe disease ((ii) and (iii) in Fig. 1). Our search terms were:

(i) (*pathogen name* OR *disease name*) AND (“aerosol production” OR “exhaled breath”) AND (“severity” OR “symptoms” OR “LRT”)

(ii)-(iii) (*pathogen name* OR *disease name*) AND (“aerosol” OR “LRT”) AND (“infection” OR “inoculation”) AND (“severity” OR “symptoms”)

Because this review covers a breadth of pathogens, as well as literature from a range of disciplines and using multiple study types, there were inconsistencies between studies in the language used to describe similar phenomena. For example, studies investigating a relationship between dose and severity for measles tended to discuss “intensity of exposure” with limited, if any, mention of infectious dose. This is likely a product of most studies being household or outbreak studies which were considering if overcrowding was a risk factor. Conversely, for influenza, the majority of studies surrounding dose-severity relationships are human challenge studies in which the dose can be measured, and hence, the dose is typically discussed explicitly. Our search terms were chosen based on the language used in our preliminary search of the literature with the aim of uncovering studies investigating the relevant relationships using a range of study techniques and terminologies.

To look for additional relevant studies, we carried out a manual search of all studies cited within these studies. All studies’ titles and abstracts were then assessed for relevance to any part of the two symptom propagation mechanisms. Overall, we included 225 studies in the review.

## 4 Biological evidence

In this section, we first provide a pathogen-agnostic overview of the evidence base related to infectious particle size and its implications on the viability of symptom propagation mechanisms relationships (Section 4.1). Our scoping literature review for 14 respiratory pathogens resulted in us observing several general relationships, independent of the pathogen, which we describe in Section 4.2.

We then demonstrate the breadth of our pathogen-specific findings, and summarise the evidence of symptom propagation for four key pathogens: influenza virus (Section 4.3), measles virus (Section 4.4), SARS-CoV-2 (Section 4.5) and *Yersinia pestis* (Section 4.6). These pathogens were chosen either due to their importance as public health threats (influenza and SARS-CoV-2) or due to them demonstrating strong evidence for symptom propagation via one of the two mechanisms (measles and *Yersinia pestis*). Our summaries of evidence of symptom propagation for each pathogen encompass both experimental studies (human volunteer challenge studies and animal model studies), hospital- and community-based studies and modelling studies (e.g. within-host immune models). We adopt a formulaic structure of: (i) introduction to the public health burden of the pathogen and discussion of general evidence of any phenomenon that could be explained by symptom propagation (e.g. clusters of severe cases); (ii) Dose-response relationship - evidence for/against a relationship between: symptom severity and pathogen load (Fig. 1(a)(i)); pathogen load and the infectious dose in secondary cases (Fig. 1(a)(ii)); a higher infectious dose and severe symptoms (Fig. 1(a)(iii)); (iii) Route-severity relationship - evidence for/against more severe symptoms increasing the likelihood of aerosolised transmission (Fig. 1(b)(i)) and infection via aerosols being more likely to cause severe symptoms than infection via other routes (Fig. 1(b)(ii) and (iii)).

Our analysis of the ten remaining pathogens - Adenovirus, *Bordetella pertussis*, Group A streptococci, MERS-CoV, *Mycobacterium tuberculosis*, RSV, Rhinovirus, SARS-CoV-1, Variola virus (Smallpox), Varicella zoster virus (chickenpox) - is provided in the Supplementary Material (File S1). A summary table contains the included studies for all 14 pathogens (see Supplementary Material, File S2).

### 4.1 Existing evidence for role of particle size

Independent of the respiratory pathogen, studies suggest that the majority of small aerosols produced during breathing originate from the LRT, being released when small airways in the lungs are opened at the end of exhalation [22–24]. In contrast, it is suggested large droplets originate from the URT because their production correlates with airflow at the beginning of exhalation [22]. Johnson *et al.* [30] found that speech and coughs produced particles in a range of sizes, with aerosols originating from the LRT and large droplets originating from the URT. Similarly, Morawska *et al.* [31] found that speech and coughs generally produced large droplets originating from the URT. Therefore, those with infection concentrated in the LRT are expected to generate larger volumes of infectious aerosols than those with infection predominantly in the URT. However, it may be possible for some amount of aerosols produced during talking and singing to originate from the URT [32].

Studies have also shown that particle size determines the potential sites of deposition because larger particles are more likely to contract the respiratory tract earlier on and are too large to enter the small airways. URT deposition has been found to increase with particle size [25], with negligible levels for droplets sized between 1 *−* 2*µm*, rising to close to 100% for droplets larger than 10*µm* [33]. In contrast, deposition in the LRT increases as particle size decreases [25, 26].

As a collective, these studies provide evidence to support route-severity relationships through the nature of aerosols and their mechanics.

### 4.2 General observations across pathogens

From our assessment of all 14 pathogens considered in our study, there was large heterogeneity between pathogens in the number of relevant studies found, ranging from 3 (for Varicella zoster virus) to 37 (for SARS-CoV-2) (Fig. 2). We summarise five general traits that were not strongly linked to a sole pathogen.

**Fig. 2.**
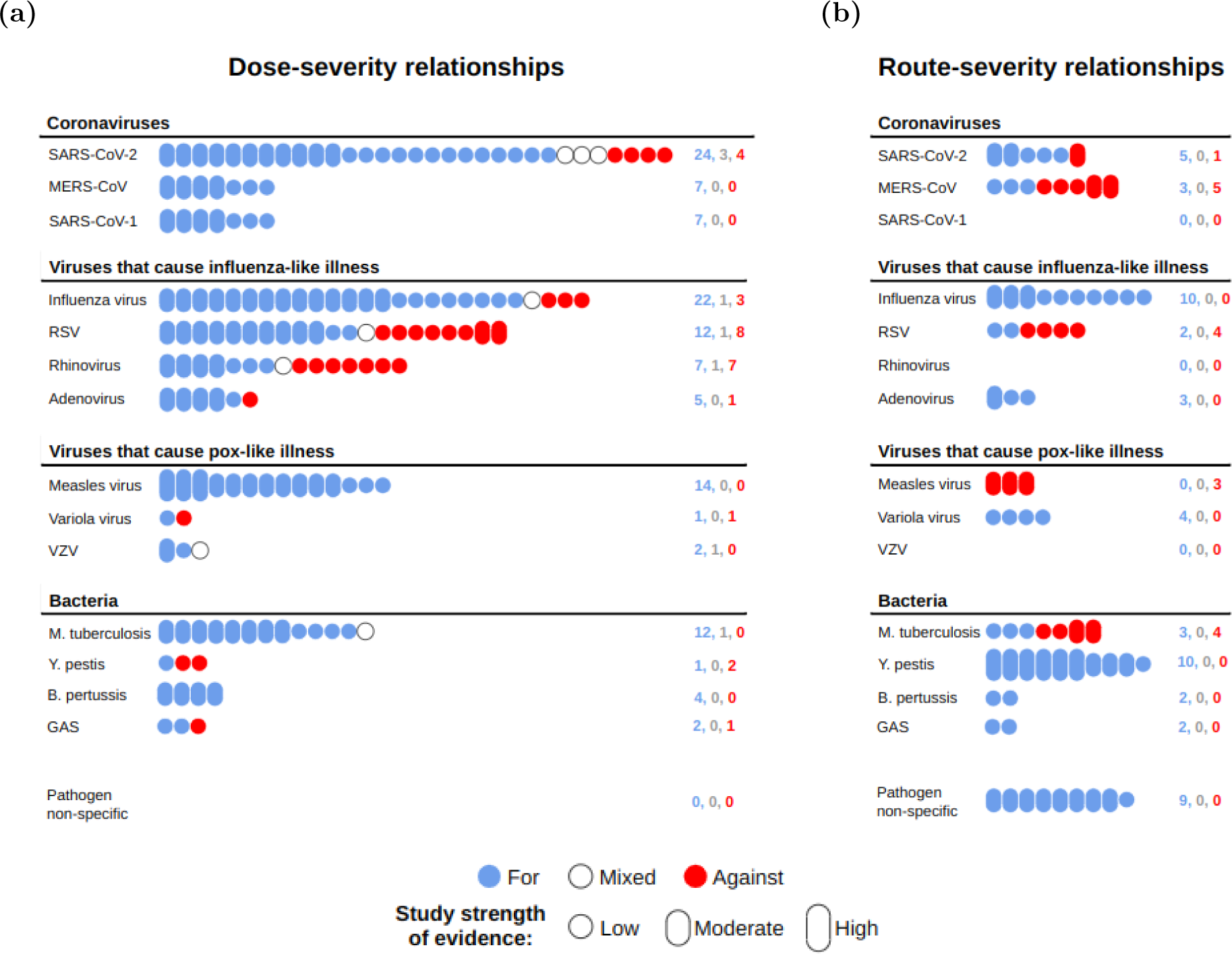
Infographic depicting the number of relevant studies found for each pathogen for the two symptom propagation mechanisms. The number of relevant studies found for each pathogen relating to (a) dose-severity relationships or (b) route-severity relationships. Colour denotes whether the study was supportive (blue) or against (red) the hypothesis, with mixed studies (white) containing findings that were both for and against or not clearly either (counts of the numbers of papers in each category are provided on the right). Bubble height denotes our classification of strength of evidence: high - a study directly investigating symptom propagation with significant findings; moderate - a study strongly related to part of the mechanism with significant findings; low - a study with either non-significant findings, or that is more weakly related to part of the mechanism. All studies are listed in the summary tables with their corresponding strength of evidence ratings (see Supplementary Material File S2). Pathogens are grouped by the type.

First, for almost all pathogens included in this study (12/14, all except RSV and rhinovirus), we found convincing evidence that symptom propagation occurs through at least one of two mechanisms: dose-severity relationships or route-severity relationships.

Second, for some pathogens, such as influenza, we found strong evidence that symptom propagation occurs through both mechanisms. For others, there was evidence for both mechanisms, but one was much more convincing; in the case of SARS-CoV-2, we could not rule out a route-severity relationship [34–38] but found notably stronger evidence in favour of a dose-severity relationship [28, 39–48].

Third, in certain pathogens, we found strong evidence for one mechanism but no evidence for the other. For measles virus, we found convincing evidence that symptom propagation occurs through a dose-severity relationship, with multiple studies finding significant correlations in severity or mortality between index and secondary cases [49–51]. However, it is unlikely that a route-severity relationship contributes to symptom propagation because measles virus is thought to not initiate infection in the URT [52–54]. These findings were echoed across other pathogens in our supplementary analysis. MERS-CoV and Mycobacterium tuberculosis are thought to predominantly initiate infection in the LRT, with initial URT infection occurring rarely, if at all [55, 56]. In contrast, RSV is thought to predominantly initiate infection in the URT, but not the LRT [57–59]. In these cases, a route-severity relationship would be unlikely to play a meaningful role in symptom propagation.

Fourth, we generally found that there were more studies relating to dose-severity relationships than route-severity relationships across pathogens. We conjecture this is due to pathogen load being easier to measure than transmission routes [60]. The evidence relating to dose-severity relationships was also more consistently in favour than those concerning route-severity relationships. The exceptions were *Yersinia pestis* and RSV, where studies suggest that a larger pathogen dose may be protective due to prompting a stronger immune response, thus resulting in reduced disease severity [61, 62].

Lastly, we found substantial evidence, not specific to any pathogen, that a route-severity relationship would be likely to occur assuming i) the pathogen can initiate infection in both the URT and LRT, and ii) the pathogen can be transmitted via aerosols and via close contact (i.e. large droplets or direct contact). This is due to aerosols released during breathing originating from the LRT and aerosols depositing in the LRT [22–26, 30]. However, these two assumptions are not necessarily sufficient. For example, for SARS-CoV-2, we know that infection can be initiated in the URT and LRT [63, 64] and we know that it can be transmitted through aerosols and close contact [65, 66], but studies found that those with asymptomatic or minimally symptomatic infection can produce aerosols to the same or even potentially a greater extent than those with moderate to severe symptoms [32, 48]. Further research is required to determine what factors are associated with increased aerosol production.

### 4.3 Influenza virus

Influenza viruses are highly transmissible and cause disease with a wide spectrum of symptoms, from mild “cold-like” symptoms to potentially fatal LRT infection [12]. There are four antigenic types of influenza: A, B, C and D. Types A and B cause the majority of infections in humans [67] and result in widespread seasonal epidemics [68]. Influenza A is capable of causing pandemics, which typically lead to elevated mortality compared to seasonal epidemics [69, 70].

#### Dose-severity relationship

We found strong evidence that those with more severe symptoms have a higher viral load, with most studies (14/15) [71–84] finding a significant relationship with one or more severity measures. However, two of these studies found a mix of significant and non-significant results [82, 83]. The remaining study [85] found a non-significant negative association. We identified two studies [84, 86] investigating whether individuals with a higher viral load infect others with a larger infectious dose, of which only one [86] found an association. In relation to a larger infectious dose causing more severe symptoms, most studies (8/9) [87–94] found an association between dose and severity, whilst one [95] found a non-significant relationship between dose and symptomatic infection, but a significant negative relationship between dose and fever.

#### More severe cases have a higher pathogen load

Hospital-based studies have found a significant relationship between URT viral load and the presence of fever [71, 72], hospitalisation [73–76], symptom score [74, 84], respiratory failure [77], abnormal findings on chest X-ray (but not worse prognosis) [78] and mortality [79]. Challenge studies have also found a significant relationship between symptom severity and viral load in humans [80] and mice [81]. However, other studies have found less consistent results. Spencer *et al.* [82] performed an analysis of 2,466 influenza-infected individuals (1,660 with influenza A and 806 with influenza B) from the US Influenza Vaccine Effectiveness Network. They found that high URT viral load (indicated by a Ct value *≤* 23) was significantly associated with self-rated illness severity for influenza A. However, for influenza B, the association between high viral load and self-rated severity was only significant when comparing severe disease against mild disease (OR 1.92; 95% CI 1.07-3.45), not when comparing very severe disease (OR 1.21; 95% CI 0.59-2.48) or moderate disease (OR 1.48; 95% CI 0.82-2.68) against mild disease. Rodrigues Guimarães Alves *et al.* [83] found that symptomatic outpatients (n=71) had significantly higher URT viral load than asymptomatic patients (*n* = 15) but found no significant difference between the viral load of symptomatic outpatients and hospitalised patients (*n* = 76). Lastly, To *et al.* [85] found that although those with fatal disease (*n* = 18) had the slowest decline in viral load, they had a lower initial URT viral load than both non-fatal severe cases (defined by the development of acute respiratory distress syndrome, *n* = 10) and mild cases (*n* = 29). However, the differences in initial viral load quantities between the case severity groups were minimal.

#### Inconclusive evidence that higher pathogen load results in infecting others with a larger dose

In a community-based study of 142 individuals, +Yan *et al.* [84] found that nasopharyngeal viral load was not a significant predictor of viral load in either large droplets (*p* = 0.48) or aerosols (*p* = 0.16). However, in an animal model study, Koster *et al.* [86] found that no transmission occurred (0/3 infected) from two ferrets with low viral load, compared to transmission consistently occurring (3/3 infected) after contact with two high viral load ferrets. Although not statistically significant, the finding is suggestive of a link between URT viral load and transmissibility. Unexpectedly, they found that viral load in exhaled aerosols was not associated with transmissibility despite aerosols being the only possible transmission route.

#### Infection with a larger dose results in more severe symptoms

Animal model studies have consistently found a relationship between inoculant dose and severity in both mice [87, 88] and ferrets [89]. Human challenge studies have also found that symptom severity increased with dose [90, 91]. In contrast, a meta-analysis across 56 volunteer challenge studies found no significant relationship between the inoculant dose and symptomatic infection (*p* = 0.12) and even found a significant negative correlation between the inoculant dose and presence of fever (OR = 0.56, 95% CI: 0.42-0.73) [95]. Handel *et al.* [92] used data from animal and human challenge studies to fit a within-host infection model and found that morbidity monotonically increased with inoculant viral load. This finding aligns with other within-host immune models: below a threshold value of initial viral load, severity is constant with respect to initial viral load, whereas above the threshold value, severity monotonically increases with initial viral load [93, 94].

#### Route-severity relationship

Influenza is widely accepted to spread through both close contact [20, 96–98] and aerosol transmission [3, 20, 96, 99–102]. Our identified relevant studies (3/3) [84, 103, 104] indicate that more severe symptoms were associated with increased aerosol production. Similarly, all studies (7/7) [95, 105–110] gave evidence that infection via aerosols resulted in more severe symptoms than infection via direct contact (including fomite transmission) or large droplet transmission.

We acknowledge that there may be variations between influenza strains that are not accounted for in these findings. For example, Kuiken *et al.* [111] found that, whilst influenza A H1N1 subtype viruses readily infect both the URT and LRT, strains within the influenza A H5N1 subtype may have a limited ability to infect the URT. To date, however, our view is there is insufficient literature to perform our analysis at a strain or subtype-specific level.

#### More severe symptoms result in increased aerosol production

Aerosols can be produced both whilst coughing and breathing [22, 84, 103, 112, 113]. If a cough is present, aerosols are produced more frequently [103] and in a larger volume [84, 104]. Despite the correlation between aerosol production and cough, it is likely that the majority of aerosols produced come from breaths, due to their greater frequency; indeed, Fabian *et al.* [112] estimate that 87% of aerosols are produced whilst breathing. Regardless, there is still likely a correlation between aerosol production and severity, due to those with severe symptoms tending to have LRT symptoms [85]. Indeed, Bischoff *et al.* [104] found that those who reported severe symptoms were significantly more likely to produce influenza aerosols. Similarly, in a community-based study, Yan *et al.* [84] concluded that URT and LRT infection occur independently and that the detection of infectious aerosols reflects infection in the LRT after finding no association between nasal shedding and aerosol production in a study of 142 symptomatic college students with confirmed influenza infection.

#### Infection via aerosols is more likely to result in severe symptoms

Animal model studies have shown that, relative to intranasal inoculation, aerosol inoculation results in more frequent LRT symptoms [105] and worse severity [106]. Mooij *et al.* [107] found more severe symptoms in macaques when inoculation was directly into the lungs (4/12 were fatal) compared to other studies that used intratracheal inoculation (generally only mild symptoms despite similarly pathogenic strains and comparable doses). Similarly, Yetter *et al.* [108] found more severe symptoms in mice when inoculation occurred in the LRT (15/16 were fatal) compared to the URT (1/16 was fatal). As reviewed in Carrat *et al.* [95], human challenge studies have also consistently found that intranasal inoculation results in mild symptoms, whereas in Alford *et al.* [109], inoculation via aerosols readily resulted in severe symptoms. To date, no further studies have been performed using aerosol inoculation. Cowling *et al.* [110] used data from randomised control trials of face masks and hand hygiene measures within 782 households to parameterise a mathematical model that accounted for three modes of transmission: aerosol, large droplet and direct contact. They inferred that the risk of fever and cough when infected via the aerosol route was around twice as high compared to infection via large droplet or direct contact routes.

### 4.4 Measles virus

Measles (also known as rubeola) is caused by the highly contagious virus of the same name. It can be seriously harmful to human health, particularly for young children [114]. The MMR (measles, mumps and rubella) vaccine protects against measles (around 96% efficacy after two doses [115]) and has been distributed widely, with over 500 million doses being administered since its introduction [116]. With the development of a highly efficacious vaccine, measles has been targeted for elimination [117]; as of 2022, elimination had been achieved in 83 countries, although elimination status had since been lost in nine of those countries [118]. In the UK, measles was initially declared eliminated in 2016, with the status then lost in 2018 and subsequently regained in 2021 [119].

There is general evidence, without specific reference to mechanisms, that symptom propagation occurs for measles. Aaby [49] found that individuals infected by someone with severe measles (indicated by pneumonia) were more likely to have severe symptoms (OR 2.90; 95% CI 1.63-5.17), and their symptoms were more likely to be fatal (OR 3.87; 95% CI 1.65-9.08). Similarly, Aaby and Leeuwenburg [50] found that the case fatality rate (CFR) was higher among cases infected by a fatal index case (OR 4.69; 95% CI 1.64-13.41) and Samb [51] found that infection by an individual with respiratory complications was more likely to result in respiratory complications in secondary cases. Based on these findings, Aaby [120] hypothesised a dose-severity relationship resulting in “feedback loops” where mild cases generate mild cases and severe cases generate severe cases. Symptom propagation could also explain the finding that the CFR increases exponentially with generations of infection [121].

#### Dose-severity relationships

We found no studies exploring whether those with more severe symptoms have higher viral loads. However, breakthrough infections (infections in those who have been vaccinated) have been shown to have a lower viral load and less severe symptoms than infected individuals who were not vaccinated [122]. We found no studies exploring whether a higher viral load was associated with infecting others with a larger infectious dose. In contrast, the effect of a larger infectious dose or increased intensity of exposure on symptom severity has been explored systematically. All included studies (13/13) [49–51, 120, 121, 123–130] found a larger infecting dose was associated with more severe symptoms. However, in one study [129] the findings were non-significant and in two studies [51, 130] they did not test for significance.

#### Infection with a larger dose results in more severe symptoms

A number of studies have found a significant increase in CFR in individuals infected within the household compared to index cases [50, 121, 123–125], suggesting an effect of increased intensity of exposure. More generally, studies have found that clustered or multiple cases (where multiple individuals within the same house are infected) have a significantly higher CFR than single cases [120, 126–128], and have suggested that this is due to increased intensity of exposure leading to larger infectious doses. Aaby *et al.* [129] found a similar correlation with no (0/24) single cases being fatal compared to 13% (10/76) for multiple cases; however, the correlation was not significant (*p* = 0.06). Aaby and Leeuwenburg [50] found that those exposed to two or more index cases had higher mortality (5/37, 14%) than those exposed to a single index case (18/303, 6%), but their finding was not significant (OR 2.47; 95% CI 0.93-6.56). However, a later hospital-based study of 221 patients found a significant relationship (OR 1.90; 95% CI 1.12-3.22) [49]. Samb [51] found that vaccinated cases produced less severe symptoms in those they infected, possibly due to their lower viral load [122]. These findings are supported by an animal model study that found higher mortality in mice that received a larger inoculant dose [130].

#### Route-severity relationships

It is generally accepted that measles is predominantly transmitted via aerosols [13, 131, 132]. However, it is unlikely that the transmission route has a direct impact on measles severity. Evidence suggests that initial infection can only occur in the LRT, with URT infection only occurring after virus is detected in the bloodstream [52–54]. In this case, symptom propagation through a route-severity relationship would not be possible, as the initial site of infection would be the LRT, regardless of the severity of the infector.

### 4.5 SARS-CoV-2 (COVID-19)

SARS-CoV-2 is the causative agent of COVID-19 and was responsible for causing a pandemic from 2020 to 2023. During this time, there were over 700 million confirmed cases worldwide and over 6 million deaths [133]. The global proliferation of SARS-CoV-2 was in part due to the large proportion of cases that were asymptomatic and their relatively high potential for onward infection, although the extent to which they contributed to transmission is still unclear [134]. This has meant that many studies have distinguished between asymptomatic and symptomatic cases when studies of other pathogens would perhaps instead compare mild and severe cases.

Throughout the COVID-19 pandemic, many variants emerged which were predominant either in particular regions or globally at certain times. There are notable differences between these variants, particularly in how transmissible they are [135] and the extent to which they evade immune responses [136]. In addition, there are differences in how readily they can transmit through aerosols and infect the LRT; for example, omicron has been found to be more likely to infect the URT than other variants [137, 138]. These factors play a role in symptom propagation, and thus we expect that the extent to which symptom propagation occurs varies between variants. To date, however, there are insufficient studies to perform our analysis at a variant-specific level.

Certain findings relating to SARS-CoV-2 could be explained by symptom propagation. For example, Guallar *et al.* [139] reported clusters of mild and severe cases during an outbreak in Madrid. In addition, Beldomenico [140] found that the case fatality ratio (CFR) was lower in countries with slow spread, leading them to suggest that high CFRs were associated with rapid transmission as a result of chains of highly infectious individuals, whose symptoms may have been more severe.

#### Dose-severity relationship

A dose-severity relationship has previously been suggested for SARS-CoV-2 in a review by Van Damme *et al.* [4]. These authors find evidence that an individual’s symptom severity is dependent on the infectious dose. They postulate that this relationship could lead to chains or clusters of severe and mild cases. Similarly, in their review, Beldomenico [140] give evidence for correlations in viral load within chains of infection, and suggest that that infection from highly infectious individuals could be more likely to be highly infectious themselves. However, they did not comment on the implications for symptom severity.

A large number of studies have explored whether those with more severe symptoms have higher viral loads. Here we discuss the findings from eight review papers [28, 39–44, 141]. All of the reviews of studies comparing severe symptoms to mild symptoms (7/7) [28, 39–44] found that viral load was significantly higher in severe cases. However, four studies [40, 42, 43, 141] reviewed comparisons between symptomatic and asymptomatic cases and found mixed results. We found three studies [45, 46, 142] comparing viral loads between moderate and mildly symptomatic patients. Of these, two [45, 46] found a significant correlation.

Although a higher viral load would likely lead to infecting others with a larger infectious dose, this idea has not been explored in depth in the literature. We found two studies comparing URT viral load to exhaled breath condensate (EBC) viral load [47, 143], but only one [47] found a correlation. Three studies [35, 48, 144] investigated whether more severe symptoms were associated with increased EBC viral load. All found a positive correlation, but in two [35, 144] the findings were not significant.

Most animal model studies (5/6) [145–149] found that an increased inoculant dose was associated with increased symptom severity; however, one [150] found no association. Most identified studies (6/7, two hospital-based studies [151, 152] and four non-pharmaceutical intervention-related studies [153–156]) found that increased intensity of exposure was associated with increased symptom severity, however one household-based study [157] found no relationship.

#### More severe cases have a higher pathogen load

Many studies have explored the relationship between viral load and severity for SARS-CoV-2. These studies predominantly use URT samples. Studies using LRT samples are not uncommon but are insufficient in number to have been reviewed separately. As such, the reviews discussed below consider URT and LRT viral load together (in addition to viral loads from other samples such as serum).

Reviews have found an often significant relationship between viral load on admission and mortality [39, 41]. They have also suggested that hospitalised patients with severe symptoms have a significantly higher viral load than those with mild symptoms [28, 39–44]. On the other hand, many reviews found mixed results regarding viral loads in symptomatic and asymptomatic patients, finding both studies where asymptomatic individuals had lower viral load and studies where viral loads were similar [40, 42, 43, 141, 158, 159]. However, these studies explore the relationship between viral load and severity at two extreme ends of the severity spectrum. There has been limited exploration of the difference in viral load between those with mild URT symptoms and those with more severe (but not hospitalised) LRT symptoms. One key exception is the study by Puchinger *et al.* [45], which found that moderate symptomatic cases (defined via the WHO symptom score, *n* = 25) had a significantly higher viral load than asymptomatic (*n* = 6) or mildly symptomatic individuals (*n* = 20*, p* = 0.01). However, Caplan *et al.* [142] found outpatients with moderate symptoms (*n* = 9) (where moderate is defined by having shortness of breath) did not have significantly higher URT viral loads than those with mild symptoms (*n* = 16*, p* = 0.24). Whilst it is clear that there is a relationship between viral load and severity in certain settings, further research is required to determine the extent of the relationship for those with moderate LRT symptoms.

#### A higher pathogen load results in infecting others with a larger dose

A few studies have attempted to determine whether increased URT viral load is associated with increased viral load in exhaled breath condensate (EBC) (and therefore with the inoculant dose in those infected); the results so far have been inconclusive. In a human challenge study, Zhou *et al.* [48] found that both nose and throat viral load significantly correlated with facemask sample viral load and Johnson *et al.* [47] found a positive correlation between EBC and URT viral load (*r* = 0.5). However, Malik *et al.* [143] found no correlation (correlation coefficient *R*^2^ *<* 0.01). It has been suggested that EBC viral load and URT viral load may not correlate due to aerosols originating from the LRT [143]. To date, no studies have compared EBC viral load with LRT viral load; this may be due to LRT viral load being more challenging to measure than URT viral load [160, 161].

Studies have begun to explore whether those with more severe symptoms have a higher EBC viral load. Sawano *et al.* [35] found that higher EBC viral load was significantly associated with the need for mechanical ventilation (*n* = 50, *p <* 0.05). They also found a positive association between EBC viral load and the need for oxygen administration and shortness of breath, but these results were not significant (*p* = 0.12 and *p* = 0.06, respectively). In a later study, Sawano *et al.* [144] again found a non-significant correlation between EBC viral load and the need for oxygen administration (*n* = 41, *p* = 0.18).

#### Infection with a larger dose results in more severe symptoms

Several animal model studies found an increase in mortality and morbidity with increasing inoculant dose [145–149]. However, Rosenke *et al.* [150] found that, although initially symptoms were more severe in hamsters given a larger inoculant dose, by day five, hamsters given the lower dose had more severe symptoms. In addition, a household contact study found no relationship between the viral load of the index case and the severity of secondary cases [157]. This result contrasts the findings of Marks *et al.* [162] who showed that individuals in contact with a high URT viral load case were significantly more likely to become symptomatic (hazard ratio per log10 increase in viral load 1·12; *p* = 0.0006). Raoult *et al.* [163] suggested that a relationship between dose and severity could be due to a larger inoculant dose overwhelming the host’s defence and after frequently detecting a state of immunosuppression hospitalised patients. Kikkert [164] instead suggested that such a relationship could be due to the initial immune response being insufficient to clear a high dose, leading to the use of a second line of defence which triggers increased inflammation.

Studies give evidence for increased intensity of exposure being associated with increased symptom severity. Maltezou *et al.* [151] found that healthcare workers with high-risk exposure (close contact with a COVID-19 case with neither party wearing a mask) were significantly more likely than those with moderate- or low-risk exposures to develop symptoms (31.9%, 22.6%, and 15.8%, respectively; *p <* 0.001) and to be hospitalised (0.8%, 0.4%, and 0.1%, respectively; *p <* 0.001). In addition, Zhang *et al.* [152] found that healthcare workers who had performed high-risk procedures, such as tracheal intubation, were significantly more likely to have their infection be symptomatic (OR 4.057; *p* = 0.026) and that healthcare workers who consistently wore respirators were significantly less likely to have their infection be symptomatic (OR 0.369; *p* = 0.001).

Dose-severity relationships have also been discussed in the context of non-pharmaceutical interventions (NPIs) such as mask-wearing and social distancing. Several studies found that, when NPIs were used, the proportion of cases that were symptomatic was greatly reduced. In military barracks, an outbreak that started before social distancing measures were introduced had 102 symptomatic cases and 113 confirmed infections by PCR test; however, in another barracks where individuals only became infected after social distancing measures were introduced, none had symptomatic infection despite 13 testing positive [153]. In a study of an outbreak in Spain, Soriano *et al.* [154] found that the proportion of individuals who tested positive by PCR test who were symptomatic was notably higher in the first wave (34/122, 27.8%) than in the second wave (5/47, 10.6%) when NPIs such as social distancing and mask-wearing were used. In their review, Gandhi and Rutherford [155] discussed three outbreak studies during which universal masking was implemented; in each case, over 80% of cases were asymptomatic. This reduction in the proportion of cases that are symptomatic has been suggested to be due to a reduction in the inoculum dose of those infected [7, 165–167]. Chan *et al.* [156] investigated the impact of mask-wearing in an animal model study involving 27 Syrian hamsters and found that the presence of a mask-like barrier not only reduced transmission but also led to reduced symptom severity in the animals that were infected.

#### Route-severity relationships

Transmission routes have been a matter of debate for SARS-CoV-2. When intervention guidelines were initially issued in 2020, most focused on close contact transmission and did not mention an airborne route [168]. Since then, many reviews have emerged suggesting that aerosol transmission had been overlooked and is, in fact, the primary transmission route [6, 66, 169, 170].

We found two studies [34, 35] showing that individuals with more severe symptoms were significantly more likely to be aerosol-positive. However, the other relevant study [48] found no relationship between symptom severity and aerosol production. All relevant studies (3/3) [36–38] found that infection via aerosols was associated with increased symptom severity.

#### More severe symptoms result in increased aerosol production

Aerosols have previously been assumed to be primarily released through coughing [171], suggesting that those with symptomatic infection are more likely to infect others via the aerosol route. However, more recent evidence has found that a substantial proportion of aerosols are produced during speaking and breathing [32, 168, 172] and studies have shown that asymptomatic individuals can transmit disease via aerosols [34, 173, 174]. However, Leding *et al.* [34] found that symptomatic individuals were significantly more likely to have SARS-CoV-2 detected in EBC than asymptomatic individuals (OR, 4.4; *p* = 0.017). In addition, Sawano *et al.* [35] found that the detection of viral RNA in EBC was significantly associated with the need for oxygen administration (*p <* 0.01), the need for mechanical ventilation (*p* = 0.04), cough (*p <* 0.01) and fever (*p* = 0.01).

More severe symptoms are associated with LRT infection. Chen *et al.* [44] found that LRT viral load was a much more accurate prognostic indicator for COVID-19 severity than URT viral load (up to 81% accuracy for LRT vs 65% for URT). Pan *et al.* [175] also found that those who had a positive facemask sample but negative URT sample had significantly higher median symptom scores than those who were facemask negative but URT positive (15 vs 3, *p* = 0.0017). Since aerosols produced during breathing originate from the LRT, LRT infection is a requirement for the production of infectious breath aerosols (see Section 4.1). Indeed, in their review, Stadnytskyi *et al.* [32] suggested that only those with clinical symptoms such as cough, reflecting LRT infection, generate aerosols during breathing. However, they suggest that speech aerosols are likely to be a predominant mode of transmission for those without symptoms because speech aerosols originate from the URT, in addition to the LRT.

Whilst evidence suggests those with more severe symptoms are more likely to be aerosol positive, this does not necessarily mean that they produce a larger number of infectious aerosols. Indeed, in a human challenge study of 36 volunteers, Zhou *et al.* [48] found that those who reported the highest symptom scores were not those who emitted the most virus.

#### Infection via aerosols is more likely to result in severe symptoms

Animal model studies have found that infection via aerosols results in more severe symptoms, compared to intranasal inoculation [36–38].

### 4.6 Yersinia pestis (Plague)

Plague, caused by the bacterium *Yersinia pestis*, predominantly comes in two forms: bubonic and pneumonic [2]. Bubonic plague is generally less severe, although it still has high mortality, and predominantly affects the lymph nodes, causing inflammation and swelling [176]. Pneumonic plague affects the LRT and has a mortality rate close to 100% when left untreated [2]. Even when initially mild or atypical cases have been reported, they still historically led to fatality [177]. However, there is some evidence that asymptomatic carriers may exist. Tieh *et al.* [177] detected *Yersinia pestis* in the throat of an otherwise healthy individual and suggested that they may be able to act as a carrier of disease. Marshall *et al.* [178] detected *Yersinia pestis* in the throats of 15 of 114 healthy people who had been in contact with an infected individual. However, if there are asymptomatic carriers of plague, they are likely to occur rarely.

#### Dose-severity relationship

Individuals with bubonic plague are generally not capable of direct human-to-human transmission; the disease is usually transmitted via a vector such as fleas or small mammals [176]. When human-to-human transmission of plague does occur, it is predominantly via the aerosol transmission route from those with pneumonic plague [2]. Those infected in this manner develop pneumonic plague [176, 179–183]. Due to pneumonic plague having an extremely high mortality rate (close to 100% if untreated [2]), current available evidence suggests there is low potential for severity to depend on infectious dose. Note, however, that the literature is inconclusive about whether asymptomatic Yersinia pestis carriers exist; such asymptomatic cases may occur as a result of a low infectious dose, but further study is required to establish this.

We found one study [184] investigating whether those with more severe symptoms had a higher bacterial load, which found a positive correlation. We found no studies exploring whether having a higher bacterial load was associated with infecting others with a larger infectious dose. On the relationship between inoculant dose and severity we found two relevant studies. One found no relationship [61] and the other found a negative relationship [62].

#### More severe cases have a higher pathogen load

In an animal model study in mice, Guinet *et al.* [184] found that bacterial loads in lymph nodes correlated with mortality.

#### Infection with a larger dose does not result in more severe symptoms

Druett *et al.* [61] found that inoculating guinea pigs with a larger dose did not lead to increased mortality in secondary cases. They did not comment on the mortality rate in the inoculated guinea pigs. Parry [62] found that rats inoculated with a larger dose had a lower mortality rate, and their deaths were delayed in fatal cases. They suggested that this was due to a large volume of inactive bacteria blocking active bacteria from potential sites of infection.

#### Route-severity relationship

Route-severity relationships are known to exist for *Yersinia pestis*. Only individuals with pneumonic plague are able to infect others through the respiratory route [2, 180–183]. Infection via this route leads to bacteria infecting the lungs, causing primary pneumonic plague in the individual infected [2, 179–183]. Individuals with pneumonic plague can also infect others via direct contact; those infected develop bubonic plague [2]. These findings are supported by Druett *et al.* [61], who found that infection of guinea pigs via large droplets resulted in URT infection, whereas aerosols lead to LRT infection. In addition, they found that mortality was four times higher in secondary cases who were in contact with aerosol-infected animals. Similarly, Agar *et al.* [185] found that rats infected via the aerosolised route developed pneumonic plague. Further, the rats were able to transmit pneumonic plague to uninoculated rats.

## 5 Symptom propagation mechanisms and infectious disease modelling

Herein we summarise the progress to date in the development of mathematical model frameworks that explicitly contain symptom propagation mechanisms (Section 5.1). To then demonstrate the importance of accounting for symptom propagation in epidemiological models, we present a case study where particular targeting of public health interventions can result in worsened epidemiological outcomes (Section 5.2).

### 5.1 Previous modelling developments

We may study the implications of symptom propagation of respiratory pathogens on epidemiological outcomes via computational simulation of an infectious disease transmission model. Here we describe the historical advancements in model frameworks towards having models that explicitly contain a symptom severity propagation action.

#### Independent strain/multi-strain type models

For respiratory pathogens, symptom severity has typically been modelled post-hoc or separately from epidemiological dynamics. For example, it has become commonplace for models to distinguish between asymptomatic and symptomatic infection, but asymptomatic infections are generally assumed to occur with a fixed probability, independent of other infected individuals [186]. An extension to this model has been explored for influenza by Paulo *et al.* [187], where the probability of severe disease depended on the proportion of the population infected at the time, although not on their severity.

#### Models capturing multi-route transmission

Other models in the literature capture multi-route transmission, but do not invoke a relationship between the route of transmission and symptom severity. For influenza, as a tool for assessing NPIs, Atkinson and Wein [188] constructed a mathematical model of aerosol and contact transmission within a single household. Another study investigated the dynamics and control of influenza under the assumptions of no, partial, or full aerosol transmission, using a model parameterised by contact network and location data [189]. Further, Cowling *et al.* [110] used data from randomised control trials of face masks and hand hygiene measures for influenza within 782 households to parameterise a mathematical model that accounted for three modes of transmission: aerosol, large droplet and direct contact.

#### Models with explicit mechanisms for the propagation of symptom severity

Embryonic attempts to incorporate symptom propagation into an epidemiological model of infectious disease transmission have been made by Ball and Britton [190, 191], Ball *et al.* [192], Earnest [193], Santermans *et al.* [194] and Harris *et al.* [195]. Ball and Britton [190] introduced the infector-dependent severity (IDS) model in which the probability of becoming severely infected was greater if infected by a severe case than if infected by a mild case. They used a stochastic epidemic model and considered the addition of two types of vaccine. This work was continued in Ball and Britton [191], where they again used the IDS model to explore vaccination, with the extension that mild cases could become severe after further exposure to disease. They found that, under this model, vaccination could lead to more people being mildly infected. In Ball *et al.* [192], the authors apply the IDS model to a household epidemic model. They determined that it would be possible to distinguish the household IDS model from a standard household model with no symptom propagation, given data on sufficiently many households.

Santermans *et al.* [194] introduced a ‘preferential model’ that works in much the same way as the IDS model, but was instead applied to a compartment SEIR infectious disease transmission model. An individual infected by an asymptomatic case was asymptomatic with probability *ϕ_a_*. If they were instead infected by a symptomatic case, they were symptomatic with probability *ϕ_s_*. This model simplifies to the so-called ‘non-preferential model’, the model with no symptom propagation, when *ϕ_s_* = 1 *− ϕ_a_*. They estimated parameters using a Markov Chain Monte Carlo (MCMC) approach applied to incidence data from 2009 H1N1 influenza pandemic. By calculating the 95% credible interval for the difference between *ϕ_s_* and 1 *− ϕ_a_*, the authors found that the preferential model did not simplify to the non-preferential model.

Earnest [193] also explored an SEIR infectious disease transmission model where the probability of mild/severe disease depended on whether the infector had mild or severe disease. Similarly, Harris *et al.* [195] studied a SARS-CoV-2 SEIR transmission model where transmission from asymptomatic (symptomatic) individuals was more likely to lead to asymptomatic (symptomatic) infection. They found that when infectious periods of asymptomatic and symptomatic infections were equal, the correlation between disease status and transmission outcomes did not affect the outbreak dynamics. In contrast, when the infectious periods of asymptomatic and symptomatic infections were dissimilar, the correlation between disease status and transmission exaggerated the effect of the difference in infectious period.

At the time of writing, models that include symptom propagation have had a similar construction, with two probabilities of having severe disease depending on whether the infector had mild or severe disease. There has been limited exploration of how the outcomes of this model differ from an analogous model without symptom propagation and a lack of sensitivity analysis to the strength of symptom propagation thus far.

### 5.2 Modelling case study

The modelling exploration of symptom propagation has been limited beyond the previously summarised results. To these prior works, we add a parsimonious mechanistic mathematical framework to model infectious disease transmission that incorporates symptom propagation of different strengths. Through application of this model, we demonstrate the importance of accounting for symptom severity propagation. Our presented example shows that, when there is a strong symptom propagation action, particular targeting of public health interventions can result in worsened epidemiological outcomes.

#### Infectious disease transmission model with symptom propagation

Taking a standard susceptible-exposed-infected-recovered (SEIR) deterministic ODE model, we supplement it with two parameters, *α* and *ν*: *α* controls the dependence of the symptom severity in the infectee on the symptom severity of the infector; *ν* sets the baseline probability of the pathogen causing severe disease in the absence of propagation effect.

Our model of symptom propagation means that an infected individual, with probability *α*, copies the symptom severity of their infector, and with probability 1 *− α* their symptom severity is assigned randomly according to the underlying probability of having severe disease, *ν* (Fig. 3).

**Fig. 3.**
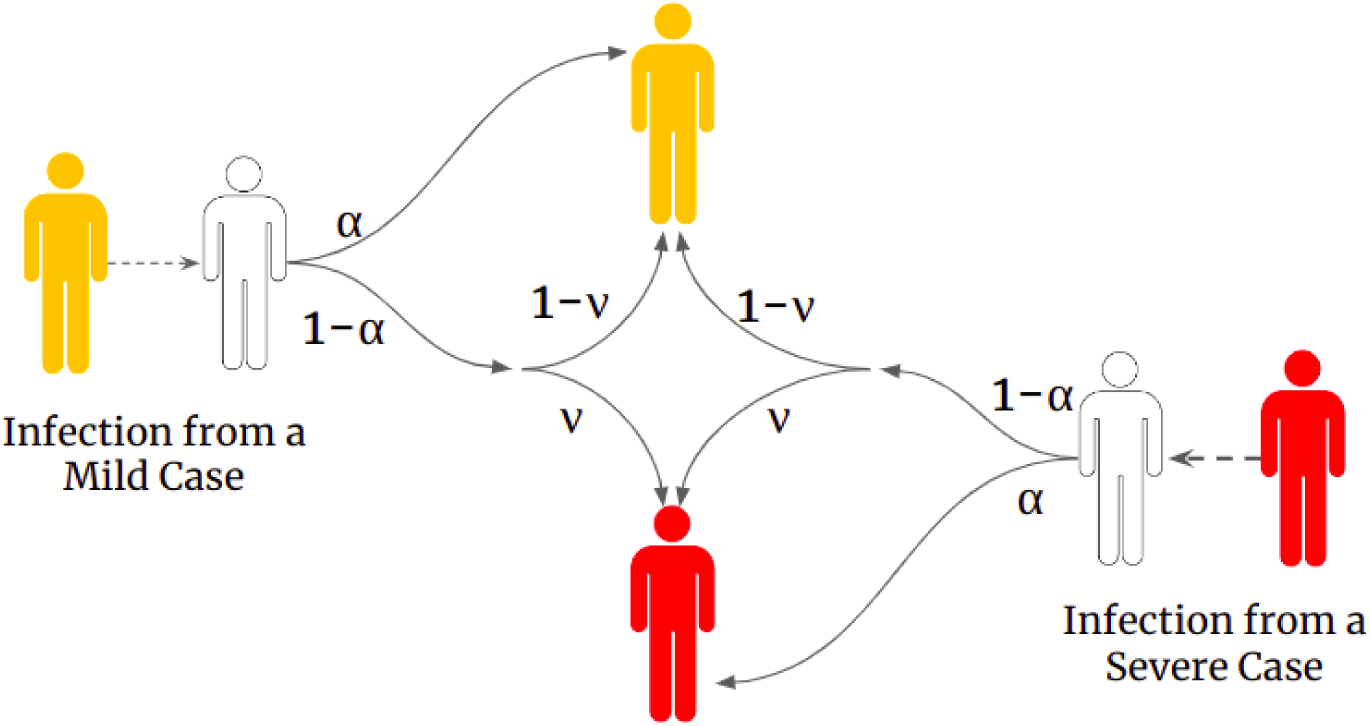
Dependence of symptom severity on *α* and *ν* (in the absence of interventions). White shaded individuals correspond to those susceptible to infection, yellow shaded individuals correspond to infectious cases with mild severity and red shaded individuals correspond to infectious cases with severe symptoms. The values on the arrows show the corresponding probability. An infected individual has probability *α* of copying the symptom severity of their infector and a probability 1 *− α* of reverting to the baseline probability of having severe disease, i.e. they develop severe disease with probability *ν*.

Mathematically, the system dynamics are characterised by the following system of ODEs:

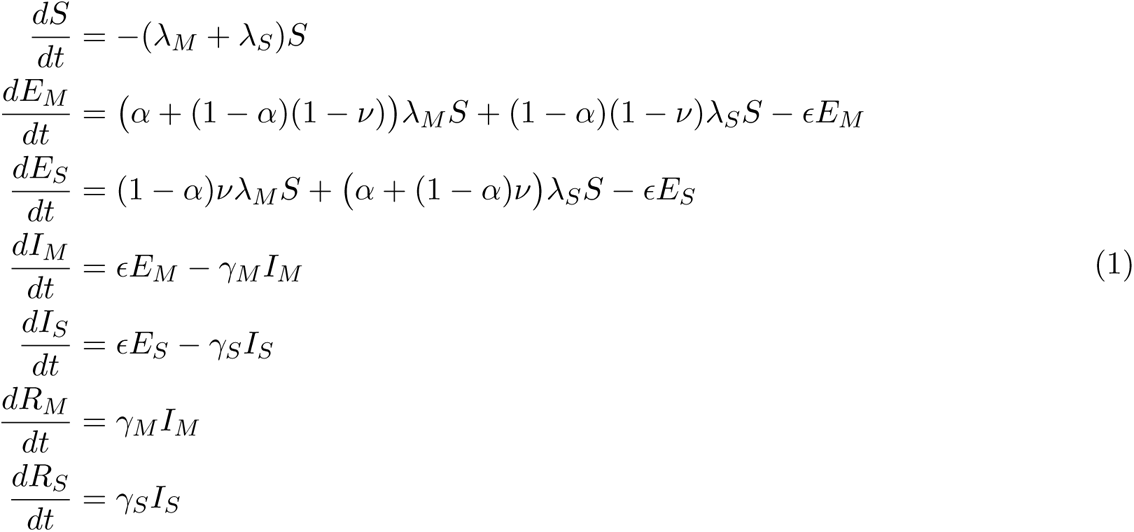

where *λ_M_* = *β_M_ I_M_* and *λ_S_* = *β_S_I_S_*.

The parameters were chosen to approximate an influenza-like pathogen (Table 1).

**Table 1.**
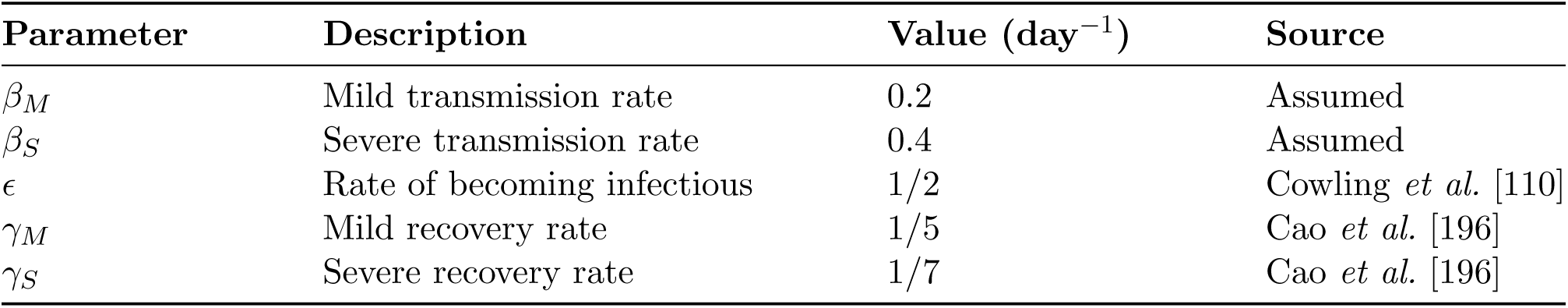
Epidemiological parameters values. The values of *β* were chosen under the assumption that severe cases were twice as transmissible as mild cases and to give a value of *R*_0_ varying between 1 to 3 for the different values of *α*and *ν*.

| Parameter | Description | Value (day<sup>-1</sup>) | Source |
| --- | --- | --- | --- |
| $\beta_M$ | Mild transmission rate | 0.2 | Assumed |
| $\beta_S$ | Severe transmission rate | 0.4 | Assumed |
| $\epsilon$ | Rate of becoming infectious | 1/2 | Cowling *et al.* [110] |
| $\gamma_M$ | Mild recovery rate | 1/5 | Cao *et al.* [196] |
| $\gamma_S$ | Severe recovery rate | 1/7 | Cao *et al.* [196] |

#### Investigating the effect of isolation measures

To evaluate the effect of symptom propagation on the effectiveness of an intervention, we introduced isolation to our model. We assumed a proportion, *Q*, of infectious individuals would be isolated. These individuals would not subsequently contribute towards onward transmission, reducing the force of infection, *λ*, by a factor of 1 *− Q*. We have assumed that symptom onset corresponds to the start of infectiousness, such that individuals are isolated as soon as they become infectious. Furthermore, we assume that the proportion of individuals isolated is independent of their symptom severity (i.e. those with mild and severe disease are equally likely to be isolated).

We considered two intervention strategies: isolating a proportion *Q* of infectious individuals and isolating a proportion *Q* of only severely infectious individuals. Denoting 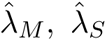 infection terms when an intervention is active, for the first intervention,

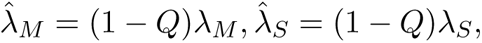

whereas for the second intervention,

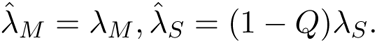

We assumed *Q* = 0.5 in each example.

We show combinations of *α* and *ν* where isolating mild infectious individuals (in addition to severe infectious individuals) would result in negative epidemiological outcomes of an increase in both severe and total cases overall (Fig. 4). Specifically, we observed an increase in total cases under sufficiently strong symptom propagation (*α >* 0.9) and almost all values of *ν* (*ν >* 0.02). Within this regime, it was notable that the percentage difference in total cases increased as *ν* decreased, reaching a maximum increase in total cases of about 10% for *α >* 0.8 and 0.02 *≤ ν ≤* 0.04. This contrasted with the dynamics for low *α*, where there was a reduction in the overall number of cases and the reductions were larger (approaching a 30% reduction) for a mid-range of *ν* (between 0.2 and 0.6).

**Fig. 4.**
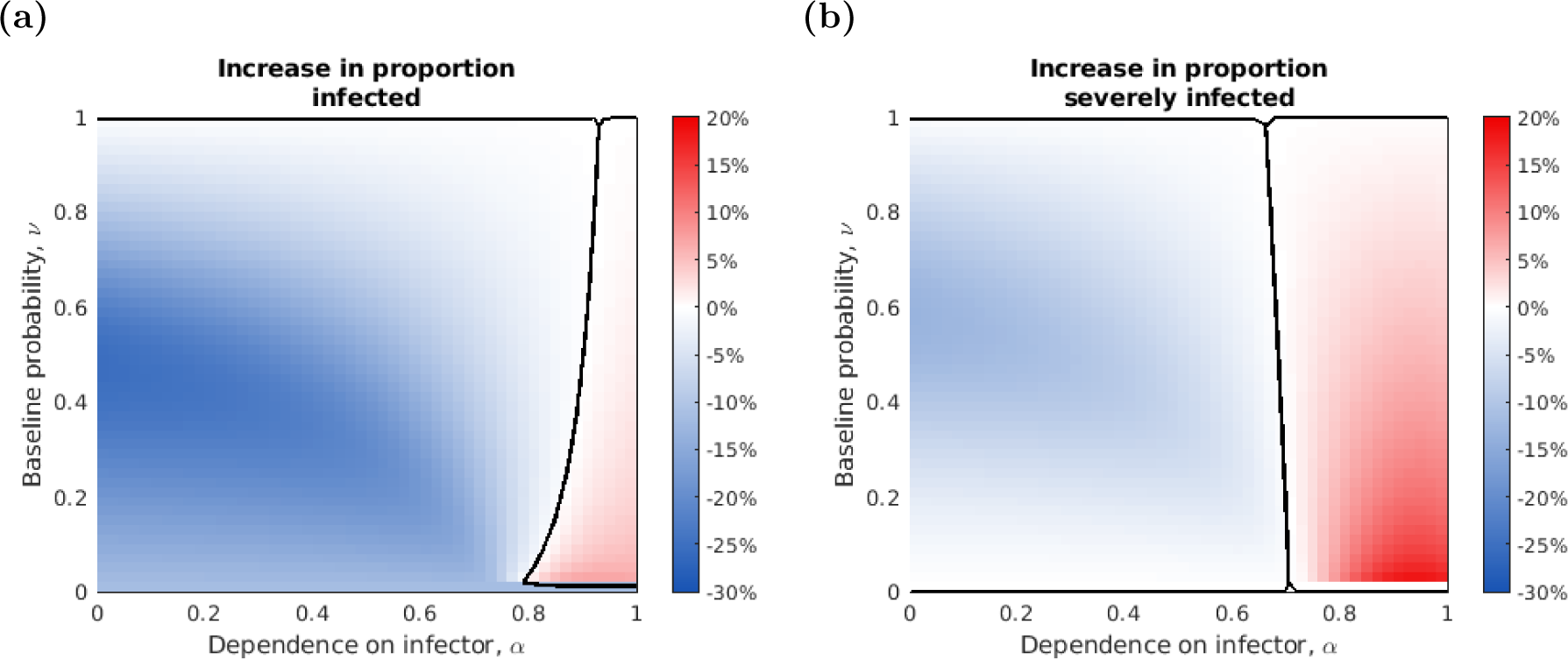
Investigating the effect of *α* and *ν* on the number of total cases and severe cases prevented by additionally isolating mild cases. (a) The difference between the overall number of cases when isolating 50% of severe cases only and the overall number of cases when isolating 50% of all cases (severe and mild). **(b)** The difference between the number of severe cases when isolating 50% of severe cases only and the number of severe cases when isolating 50% of all cases (severe and mild). Shading denotes the cases prevented (as a percentage of the population): blue denotes values where isolating mild cases decreased the number of cases; red denotes values where isolating mild cases increased the number of cases. Black solid lines represent parameter combinations where no cases were prevented by additionally isolating mild cases.

On the other hand, we found an increase in the number of severe cases when *α >* 0.7, independent of *ν*. However, there was again a trend in the magnitude of the increase and value of *ν*, with the greatest increases of above 20% being returned when *ν* was small (approximately 0.02-0.04).

Overall, we found negative implications for additionally isolating mild infectious individuals when *α >* 0.7. This effect occurs because, independent of symptom propagation, isolating only severely infected individuals increases the proportion of new infections that are generated by mildly infected individuals. Therefore, under symptom propagation, a larger proportion of newly infected individuals will have mild symptoms. Once recovered, these individuals are then removed from the susceptible pool, curtailing overall rates of transmission. Furthermore, since mild individuals are assumed to be less infectious than severe cases, the reproduction number is reduced (i.e. on average, each infectious individual produces fewer secondary infections). When symptom propagation is sufficiently strong, the effects on overall transmission of reducing the susceptible pool (which act by permitting higher rates of ongoing mild infections) begin to outweigh those of isolating mild cases (which act by reducing transmission rates). Thus, it is more effective to only isolate those with severe symptoms.

Although we only found negative overall health implications of isolating mildly infected individuals for relatively high strengths of symptom propagation, this example demonstrates the reduction in effectiveness in isolating mildly infected individuals due to symptom propagation. For medium strengths of symptom propagation (*α ≈* 0.5), isolating both mild and severely infected individuals does reduce the total number of cases, but at the cost of having to isolate many more individuals. Thus, even a relatively low strength of symptom propagation has the potential to shift the balance between isolation strategies in terms of cost-effectiveness.

Though a simplified example, we demonstrate the plausibility of particular combinations of symptom propagation mechanisms and characteristics of public health interventions resulting in unwanted epidemiological outcomes.

## 6 Discussion

Studies investigating the mechanisms behind symptom propagation have been performed for decades. For one key pathogen, *Yersinia pestis*, the causative agent of plague, the symptoms an individual experiences have been known to depend on the symptoms of their infector since the 1910s [183]. Nonetheless, symptom propagation for other pathogens of public health interest has only recently been acknowledged and is understudied. The existing literature has focused on other specialised research areas, such as dose-response relationships, aerosol and contact transmission routes and the effect of non-pharmaceutical interventions to reduce symptom severity. Our review fills a knowledge gap by collating evidence for symptom propagation for a range of respiratory pathogens. We additionally demonstrate through simulation of a mathematical model the importance of symptom propagation on intervention strategies.

For almost all pathogens in this study, we found convincing evidence that symptom propagation occurs through at least one of two mechanisms: dose-severity relationships or route-severity relationships. We also found considerable heterogeneity between pathogens in the relative importance of these two mechanisms, highlighting the importance of the individual consideration of each pathogen. Although symptom propagation in general has not previously been reviewed, our findings align with previous reviews which investigated either dose-severity or route-severity relationships. Van Damme *et al.* [4] reviewed evidence for a dose-response relationship leading to chains or clusters of severe disease for SARS-CoV-2. Milton [197] reviewed evidence for smallpox transmission routes and introduced the term “anisotropic” to describe when the transmission route alters the severity of disease. Similarly, Tellier [99] suggest a potential relationship between transmission route and disease severity for influenza. However, to date, no studies have investigated route-severity relationships in depth.

### Limitations of the biological evidence scoping review

Through conducting this study, particular limitations arose that warrant consideration when interpreting our findings. We discuss four examples. The first main limitation of our scoping review was that our analysis was not performed at a strain or subtype-specific level. We are aware that, particularly for SARS-CoV-2 and influenza, there are notable differences between strains that would impact the occurrence of symptom propagation. For example, varying pathogens loads between strains [135, 198] or differences in transmission routes or sites where the pathogen can initiate infection [137, 138]. Our view is that there is currently insufficient literature to perform our study at this refined level; indeed, for many pathogens, there is insufficient literature even at the pathogen level.

Second, there were some limitations in the studies reviewed regarding dose-severity relationships. In brief, comparison between studies is complex. Pathogen load significantly depends on how long the individual has been infected for [85, 199, 200] and the site at which the sample is taken [28, 42], and there is substantial heterogeneity across studies in both how and when pathogen load is measured. In particular, some measure pathogen load early on during infection [77, 142] or upon hospital admission [73, 83], whereas others measure peak or mean pathogen load [71, 80] or at a fixed time independent of individual patients’ duration of infection [84]. Studies may also use either URT or LRT samples [44, 201] which are not necessarily correlated [28]. For example, an individual with severe disease may have very high LRT pathogen load but low URT pathogen load [202]. In addition, despite LRT pathogen load being considered a better indicator for infection or severe disease for many pathogens [201, 203, 204], often URT measurements are taken instead as they are easier to perform [160, 161].

Another aspect relating to dose-severity relationships is that, for most pathogens, an increasing infectious dose is associated with a higher probability of successful infection [15, 146, 205]. Thus more cases are generated at a higher dose. Therefore, even in the absence of symptom propagation (i.e. when the proportion of cases that are severe is independent of dose), we expect a higher dose to generate a greater number of severe cases. This dynamic needs to be accounted for in observational studies by measuring what proportion of individuals are successfully infected to determine what proportion of infections are severe; it is not sufficient to simply compare the number of severe cases generated at different doses. Such analyses are often not performed.

The third set of limitations relates to determining the level of support for route-severity relationships. One of our key findings is that to enable ascertainment of whether route-severity relationships occur, it is crucial to determine where a pathogen can initiate infection. Determining whether infection can be initiated in the URT or LRT is quite difficult, especially because the detection of pathogens at a site is insufficient. For example, for some pathogens, initial infection of the LRT is not thought to be possible, but the pathogen can spread to the LRT after URT infection [57–59]. We also used URT and LRT infection as distinguishing factors to categorise disease severity. Whilst LRT symptoms are generally more severe than URT symptoms, the definition of severe disease varies between pathogens. In certain cases, severity is categorised by systemic symptoms like fever [206–208] or septic shock [209]. Focusing on SARS-CoV-2, there is evidence that LRT infection is not limited to those with severe disease, with even asymptomatic patients having LRT viral load detected [210]. Further research is required for multiple reasons. One is to clarify the link between severity and LRT infection. Another is to develop a more formal definition of symptom severity, thus allowing for consistent terminology across clinical, modelling and health economic disciplines. We believe these efforts would benefit from clinical input, helping devise a new framework for categorising clinical outcomes plus the formulation of associated data collection protocols.

The fourth and final limitation we discuss is that, for some pathogens, there were insufficient studies relating to a particular mechanism to conclude whether it would contribute to symptom propagation. There were even notable differences when considering the subsections of a mechanism for a specific pathogen. One notable example was for rhinovirus. We found 15 studies exploring whether those with more severe disease had a higher viral load, but no studies related to whether a higher viral load was associated with a larger infectious dose or if a larger infectious dose was associated with more severe symptoms. Our findings motivate an increased breadth of research for certain pathogens to determine more clearly the contribution of potential symptom propagation mechanisms for these pathogens.

### Mathematical modelling: A tool for discovery

There are other factors impacting severity that are challenging to account for, including pathogen strain and genetic factors. As a result, there are limitations even in studies explicitly investigating correlations in disease severity or mortality, such as those for measles [49–51]. These factors would be complicated to account for outside of a controlled human challenge study. Still, these studies would come with their issues, including ethical issues surrounding intentionally causing severe infection. With the difficulties that come with these hypothetical studies, it is imperative that we use modelling tools to supplement our current knowledge from biological studies. Modelling can be an applied tool to help determine whether and to what extent symptom propagation occurs for a given pathogen.

There has been limited exploration of symptom propagation within the modelling literature. All of the modelling studies we identified with explicit mechanisms for symptom propagation used a fixed probability of developing severe disease (or, in some cases, symptomatic disease), where this probability depended on whether the infector had mild or severe disease [190–194]. None of these studies allowed for variations in the strengths of symptom propagation. Thus, none of these prior studies explored the effect of varying the strength of symptom propagation and only compared their fixed probability model against one with no symptom propagation.

We have shown through a simple model case study that symptom propagation can have dramatic implications for the effectiveness of intervention strategies. Under strong symptom propagation, isolating mildly infected individuals (in addition to those severely infected) led to an increase in both severe and total cases, compared to only isolating severe cases. Whilst our isolation model is quite rudimentary, our findings motivate further modelling work by demonstrating potentially important public health impacts of symptom propagation. In addition to impacting isolation and test-and-trace measures, we believe that symptom propagation could increase the effectiveness of NPIs, such as mask-wearing or social distancing, which act to reduce the pathogen dose individuals are infected with.

### Outlook

We have collated the evidence on the presence or absence of symptom propagation for pathogens that have (historically and presently) inflicted substantial burden upon public health. However, when a novel pathogen with pandemic potential in humans emerges, initially there will almost certainly be a scarcity of relevant data despite health decision makers desiring the use of models and data from multiple sources [211]. A key characteristic of the novel pathogen could be symptom propagation. A key area of future study motivated by this review is the development of more realistic mathematical models of symptom propagation. Our novel mathematical framework has been developed with the ultimate aim to parameterise *α* and determine the strength of symptom propagation. Understanding whether, and to what extent, symptom propagation occurs would allow policy makers to have a more complete understanding of the impact of intervention strategies and thus have a more effective response.

However, caution is required when inferring the strength of symptom propagation from data. Symptom severity has strong effects on patterns of contact and behaviour which would need to be controlled for. In addition, non-pharmaceutical interventions such as mask-wearing or social distancing could reduce the effect of symptom propagation due to a general decrease in disease severity caused by a reduction in the infectious dose. Furthermore, there are potential confounding factors which could result in patterns that look similar to those of symptom propagation. For example, correlations in symptom severity could be due to genetic factors when individuals infect those that they are related to. Alternatively, weather could lead to correlations in symptom severity, for example, due to aerosols being more readily produced in lower humidity. These limitations highlight the importance of experimental studies, which can investigate symptom propagation in a controlled environment where these factors can be accounted for.

On the other hand, experimental studies come with their own limitations. Often sample sizes are limited due to ethical considerations, particularly in animal-model and human challenge studies. These small sample sizes can lead to a lack of statistical significance which may lead to findings being overinterpreted, or on the contrary, dismissed. Overall, our view is that multiple methods are required to determine the extent to which symptom propagation occurs, drawing evidence from both experimental and observational studies.

Future research could also extend our understanding of symptom propagation beyond correlations in symptom severity to look at the propagation of symptom sets. Our investigation of route-severity relationships has begun to uncover this type of relationship, with evidence showing that symptoms specifically associated with LRT infection can propagate. The propagation of symptom sets may have widespread implications for public health, for example if chains of atypical or less easily detectable symptoms occur that surveillance strategies could miss.

## Conclusion

In this paper, we have reviewed the epidemiological and biological evidence for the propagation of symptom severity for a broad range of respiratory pathogens of public health concern. We demonstrate how symptom propagation is a widespread phenomenon that impacts the transmission dynamics of many respiratory pathogens. There is, however, still uncertainty surrounding symptom propagation for many pathogens, motivating an expansion of our biological evidence knowledge base. These efforts can be aided by the use of modelling and robust parameter inference to determine symptom propagation-related parameters. Our presentation of a mathematical framework unifying a standard infectious disease transmission model with a symptom propagation mechanism has demonstrated how negative public health outcomes can result when symptom propagation is strongly present in the dynamics. In summary, we believe that increased awareness and study of symptom propagation will deliver crucial infectious disease insights; the downstream implications for public health policy will subsequently allow the general public to make more informed decisions to limit the transmission of severe disease within their communities.

## Author contributions

**Phoebe Asplin:** Conceptualisation, Data curation, Formal analysis, Investigation, Methodology, Software, Validation, Visualisation, Writing - Original Draft, Writing - Review & Editing.

**Rebecca Mancy:** Conceptualisation, Investigation, Methodology, Supervision, Visualisation, Writing - Review & Editing.

**Tom Finnie:** Conceptualisation, Methodology, Visualisation, Writing - Review & Editing.

**Fergus Cumming:** Conceptualisation, Methodology, Visualisation, Writing - Review & Editing.

**Matt J. Keeling:** Conceptualisation, Investigation, Methodology, Supervision, Visualisation, Writing - Review & Editing.

**Edward M. Hill:** Conceptualisation, Investigation, Methodology, Software, Supervision, Validation, Visualisation, Writing - Original Draft, Writing - Review & Editing.

## Data availability

All data utilised in this study are publicly available, with relevant references and data repositories provided.

## Code availability

The code repository for the study is available at: https://github.com/pasplin/symptom-propagation-case-study. Archived code: https://doi.org/10.5281/zenodo.10412708.

## Data Availability

All data produced are available online at https://github.com/pasplin/symptom-propagation-case-study

