## Supplementary Material S1 for "Symptom propagation in respiratory pathogens of public health concern: a review of the evidence"

### Supplementary File S1
### Symptom propagation in respiratory pathogens of public health concern: a review of the evidence

Phoebe Asplin[1,2,3*], Rebecca Mancy[4,5], Thomas Finnie[6], Fergus Cumming[7], Matt J. Keeling[2,3,8], Edward M. Hill[2,3*]

**1** EPSRC & MRC Centre for Doctoral Training in Mathematics for Real-World Systems, University of Warwick, Coventry, United Kingdom.
**2** Mathematics Institute, University of Warwick, Coventry, United Kingdom.
**3** The Zeeman Institute for Systems Biology & Infectious Disease Epidemiology Research, University of Warwick, Coventry, United Kingdom.
**4** School of Biodiversity, One Health and Veterinary Medicine, University of Glasgow, Glasgow, United Kingdom.
**5** MRC/CSO Social and Public Health Sciences Unit, University of Glasgow, Glasgow, United Kingdom.
**6** Data, Analytics and Surveillance, UK Health Security Agency, London, United Kingdom.
**7** Foreign, Commonwealth and Development Office, London, United Kingdom.
**8** School of Life Sciences, University of Warwick, Coventry, United Kingdom.

#### Table of Contents

**S1 Additional figures** 2

**S2 Biological evidence for symptom propagation in 10 additional pathogens** 3
- S2.1 Coronaviruses 3
  - S2.1.1 MERS-CoV 3
  - S2.1.2 SARS-CoV-1 3
- S2.2 Viruses that cause influenza-like illness 4
  - S2.2.1 Respiratory Syncytial Virus 4
  - S2.2.2 Rhinovirus 6
  - S2.2.3 Adenovirus 7
- S2.3 Viruses that cause pox-like illness 8
  - S2.3.1 Variola virus (Smallpox) 8
  - S2.3.2 Varicella zoster virus (Chickenpox) 9
- S2.4 Bacteria 10
  - S2.4.1 *Mycobacterium tuberculosis* 10
  - S2.4.2 *Bordetella pertussis* 12
  - S2.4.3 Group A streptococci (GAS) 13

### S1 Additional figures

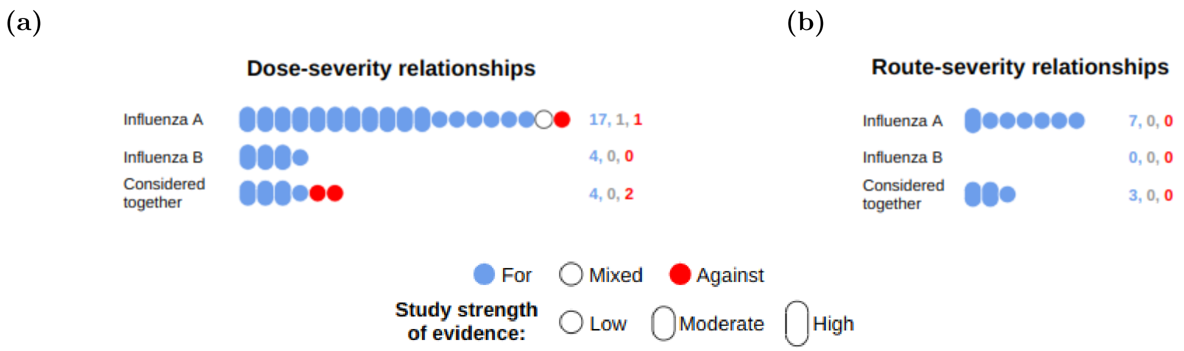


**Fig. S1. Infographic depicting the number of relevant studies found for each influenza type for the two symptom propagation mechanisms.** The number of relevant studies found for each influenza type relating to **(a)** dose-severity relationships or **(b)** route-severity relationships. Colour denotes whether the study was supportive (blue) or against (red) the hypothesis, with mixed studies (white) containing findings that were both for and against or not clearly either (counts of the numbers of papers in each category are provided on the right). Bubble height denotes our classification of strength of evidence: high - a study directly investigating symptom propagation with significant findings; moderate - a study strongly related to part of the mechanism with significant findings; low - a study with either non-significant findings, or that is more weakly related to part of the mechanism. Here, the studies listed for either Influenza A or Influenza B either only considered one of the two types or considered both independently. 'Considered together' contains studies in which both Influenza A and B were present, but the analysis did not consider them separately. All studies are listed in the summary tables with their corresponding strength of evidence ratings (see Supplementary Material File S2). Pathogens are grouped by the type.

### S2 Biological evidence for symptom propagation in 10 additional pathogens

#### S2.1 Coronaviruses

##### S2.1.1 MERS-CoV

MERS-CoV is the causative agent of MERS and was first identified in Saudi Arabia in 2012 [1]. MERS-CoV is of concern due to its high mortality rate, which has been estimated to be over 20% [2]. Although human-to-human transmission of MERS-CoV is possible, it occurs somewhat rarely, and MERS-CoV is not thought to have pandemic potential [3].

###### Dose-severity relationships

Evidence suggests a relationship between viral load and symptom severity, with all identified studies finding a significant relationship (5/5) [4–8]. We found no studies exploring whether a higher viral load was associated with infecting others with a larger infecting dose. Two studies identified [9, 10] found that a larger dose led to more severe symptoms.

*More severe cases have a higher pathogen load*

Oh *et al.* [4] found that those with severe symptoms had a significantly higher viral load in throat samples than those with mild symptoms ($p < 0.001$) and a non-significant relationship between nasopharyngeal viral load and severity ($p = 0.06$). Others have found a significant relationship between viral load and mortality [5–8].

*Infection with a larger dose results in more severe symptoms*

Animal model studies have found more severe symptoms in mice inoculated with a larger dose [9, 10].

###### Route-severity relationships

There is evidence for aerosol transmission [11, 12] for MERS. Some argue aerosol transmission is its only viable transmission route [12], due to the suggestion that infection is only possible in the LRT (supported by the finding that limited replication is possible in the URT due to a lack of DPP4 expression - the receptor used by the virus [13, 14]). This is also supported by the fact that the majority of disease symptoms are either systemic or LRT-related, but not URT-related [15]. In addition, LRT samples are recommended for MERS-CoV PCR testing, despite being more difficult to obtain [16] However, there is insufficient data to confirm whether MERS-CoV replicates solely, or even preferentially, in the LRT [17]. For example, in their review of MERS-CoV transmission and pathogenesis, de Wit *et al.* [18] found MERS-CoV replication in both the URT and LRT and Feikin *et al.* [5] found a significant association between URT viral load and severity.

##### S2.1.2 SARS-CoV-1

SARS-CoV-1 is the causative agent of SARS disease, and was first identified in 2003 during an outbreak in China [19]. This outbreak resulted in around 8,000 cases [20] and the disease was successfully contained, meaning that there have been no cases reported globally since 2004 [21]. However, SARS-CoV-1 is still circulating in wild animals and has the potential to re-emerge [22].

##### Dose-severity relationships

All studies identified (5/5) [23–27] found that those with severe symptoms had significantly higher viral loads. We found no studies exploring whether a higher viral load was associated with infecting others with a larger dose. We also found no studies directly investigating the effect of a larger infecting dose on symptom severity. However, we found two studies [28, 29] investigating whether measures of intensity of exposure were correlated with symptom severity, of which only one [29] found a positive relationship.

*More severe cases have a higher pathogen load*

Hospital-based studies have found that URT viral load was significantly associated with mortality [23–25] and acute respiratory distress syndrome [25]. Ng *et al.* [26] also found that serum viral load on admission was significantly associated with ICU admission. Hung *et al.* [27] also found that URT and serum viral load were significantly associated with diarrhea, respiratory failure and mortality. However, it should be noted that they measured viral load between 10 to 15 days after symptom onset. At this point, the median viral load was 0log10 copies/mL in patients without these severity markers, despite many of them testing positive by PCR test.

*Infection with a larger dose results in more severe symptoms*

In a hospital-based study of 45 healthcare workers, Wilder-Smith *et al.* [28] found no correlation between intensity of exposure (measured through closeness and duration of contact with a SARS patient) and the development of pneumonia. However, they did find that mask-wearing significantly reduced the probability of symptomatic infection; 3/6 (50%) of asymptomatic cases had worn a mask compared to 3/37 (8%) of cases with pneumonia. Chu *et al.* [29] performed a study of 321 infected residents of a housing estate. They found that individuals who lived in the same residential block as the index case had a higher viral load and increased mortality than those in other blocks.

##### Route-severity relationships

SARS-CoV can be transmitted through both aerosols and close contact [30–32]. The primary receptor for SARS-CoV (ACE2) is present throughout the respiratory tract [33, 34]. Thus, it is likely that SARS-CoV can initiate infection in both the URT and LRT. Furthermore, SARS disease is characterised by severe LRT symptoms [35], but it has also been detected in URT samples [36].

It is certainly possible for route-severity relationships to occur for SARS-CoV, but to date, no studies have been performed exploring whether more severe symptoms were associated with increased aerosol production, or whether infection via aerosols was associated with increased symptom severity compared to other transmission routes.

#### S2.2 Viruses that cause influenza-like illness

##### S2.2.1 Respiratory Syncytial Virus

Respiratory syncytial virus (RSV) is a common virus which generally causes mild disease [37]. However, it can cause serious disease, particularly in infants and older adults [38]. RSV is the most common cause of LRT infection in children [39].

##### Dose-severity relationship

We found several studies exploring whether those with more severe symptoms have higher viral loads. Many studies (11/16) [40–50] found that viral load was significantly correlated with one or more measure of symptom severity, although one [50] also found no association between viral load and hospitalisation. Three studies [39, 51, 52] found no significant correlation and two [53, 54] found a significant negative correlation between viral load and symptom severity. We found no studies exploring whether a higher viral load was associated with infecting others with a larger dose. Out of five studies investigating the effect of a larger infecting dose on symptom severity, two [55, 56] found a positive association, whereas the others (3/4) [41, 46, 48] found no relationship.

*Inconclusive evidence that more severe cases have a higher pathogen load*

Studies have found a significant correlation between viral load and symptom score [40–45], hospitalisation [46, 47], length of hospital stay [45, 48], respiratory failure [46, 50] and ICU admission [46, 48, 57]. Others found a non-significant correlation between viral load and any correlates of symptom severity [39], LRT infection [51], hospitalisation [57], ICU admission [40] and death [40]. However, some studies have found no relationship between measures of severity and viral load. Duncan *et al.* [50] found no relationship between viral load and hospitalisation (OR 1.0; 95% CI 0.5-1.7). Mazur *et al.* [52] found no correlation between viral load and life-threatening disease (OR 1.0; 95% CI, 0.94–1.1) or increased length of hospitalisation (OR 1.0; 95% CI, 0.98–1.0). Hijano *et al.* [40] found that the correlation between viral load and LRT infection was minimal (OR 1.09; p=0.76). Garcia-Mauriño *et al.* [53] even found that outpatients had a significantly higher initial viral load than inpatients ($p = 0.003$) and that viral load correlated inversely with severity score ($p < 0.001$). They suggested that their findings were due to a high initial viral load causing a strong, protective immune response. This hypothesis is corroborated by Piedra *et al.* [54], who found a significant correlation between initial viral load and immune response biomarkers, in addition to a significant negative correlation with symptom severity.

*Inconclusive evidence that infection with a larger dose results in more severe symptoms*

A relationship between the inoculant dose and severity has been hypothesised; Hall and Douglas [55] found less severe symptoms from fomite transmission compared to large droplet transmission, and suggested that it was due to a smaller inoculum dose. However, no study has been able to find a relationship between the intensity of exposure (measured by the number of children in a household or attendance at day care) and subsequent viral load [46, 48], and a dose-varying human challenge studies found no relationship between the inoculant dose and symptom severity [41]. [56] also performed a dose-varying human challenge study, but none of the infected individuals developed symptomatic disease. They did, however, find that those infected with a larger inoculant dose had an increased viral load.

##### Route-severity relationship

RSV is typically assumed to primarily spread through close contact transmission [58–60]. Hall and Douglas [55] found that RSV could be transmitted through either large droplets or fomite transmission, but reported no evidence for long-range aerosol transmission ($p < 0.05$). Despite this, RSV has been detected in air filters in hospitals [61–65] and in a daycare centre [66], leading some to suggest that aerosols may play a role in RSV transmission [62, 67]. It is unclear what proportion of the RSV in these air filters is as a result of aerosols, as opposed to large droplets. Whilst Kulkarni *et al.* [63] found a "substantial amount" of RSV RNA in particles $< 4.7\mu m$ in diameter, other studies found that less than 10% of RSV RNA was in particles $< 4.1\mu m$ in diameter [62, 65], compared to 42% for Influenza A [62].

Even if aerosol transmission occurs, it is unclear if RSV is capable of initiating infection in the LRT, despite LRT symptoms being a common occurrence for RSV [68]. Direct infection of the URT is well established and is predominantly assumed to be a prerequisite for LRT infection, with direct infection of the LRT not being possible [69, 70]. This suggestion has been supported by an animal model study investigating how infection develops in mice [71]. However, Johnson *et al.* [72] suggested that direct infection of the LRT can occur after finding evidence of direct infection of the alveoli during a series of autopsies. This idea has not yet been supported by other studies.

##### S2.2.2 Rhinovirus

Rhinoviruses (RhVs) are the most frequent cause of the common cold and are generally characterised by mild but widespread illness [73]. They do, however, have the ability to cause severe disease, particularly in immunocompromised people [74] and people with asthma [75].

**Dose-severity relationship**

We found mixed results regarding whether those with more severe symptoms had a higher viral load. Several papers (7/15) [76–82] found a significant relationship between severity of symptoms and viral load. Although, in two of these studies [80, 81], the correlation was only significant when the study was restricted to a subset of the population and in another [82] the correlation was weak and only significant for one out of three measures of severity. One study [83] found a positive but not significant correlation. Several studies (7/15) [39, 44, 82, 84–87] found no relationship and one study [88] found a significant negative relationship between viral load and severity, but only in coinfections.

We found no studies exploring whether a higher viral load was associated with infecting others with a larger dose. Similarly, we found no studies investigating the effect of a larger infecting dose on symptom severity.

*Inconclusive evidence that more severe cases have a higher pathogen load*

Hospital-based studies have found a significant correlation between viral load and symptom score [76, 78] and hospital length of stay [79]. Similarly, a human challenge study of 24 volunteers found that high viral load was significantly associated with self-rated severity score [77]. Message *et al.* [80] found that those with LRT symptoms had a higher viral load. The association was significant in asthmatic patients ($n = 11$) but not in non-asthmatic patients ($n = 17$). Takeyama *et al.* [81] found a significant association between viral load and severity score in children $\geq 11$ months old ($r = 0.407, p = 0.032$). The association was not significant when all patients were considered ($r = 0.126, p = 0.421$). Piralla *et al.* [83] found that high viral load was associated with pneumonia and respiratory distress, but the relationships were not significant.

Sanchez-Codez *et al.* [82] found that low viral load significantly reduced the risk of ICU admission, although the association was weak (OR 0.97; 95% CI 0.95-0.99). They also found no association between low viral load and reduced risk of hospitalisation (OR 0.99; 95% CI 0.97-1.01) or reduced risk of respiratory failure (OR 0.97; 95% CI 0.97-1.00). However, it should be noted that they did not account for the duration of illness at the time of sample collection, which could explain the lack of association - we would expect that patients in hospital or with respiratory failure are further along in their disease course, which is associated with a reduction in viral load [78, 87].

Several hospital-based studies found no association between viral load and severity score [44, 87], hospitalisation [84, 85], hospital length of stay [39, 86], respiratory failure [39] or ICU admission [84, 85]. In addition, Do *et al.* [88] found that high viral load was significantly negatively associated with severity score ($\rho = -0.09, p = 0.04$) but only in coinfections ($n = 119$), not single infections ($n = 87$). This may be due to RhV acting to reduce the severity of the other infection; Van Leuven *et al.* [89] found that infection by RhV reduces the severity of subsequent viral infections.

**Route-severity relationship**

There is evidence that RhVs have the potential to be transmitted through both the aerosol [90–94] and close contact [91–93, 95] transmission routes. Studies have found that RhV can initiate infection in both the URT and LRT [96–99], with LRT infection generally being associated with increased severity [93, 98].

However, we found no studies exploring whether more severe symptoms were associated with increased aerosol production. We also found no studies directly investigating whether infection via aerosols was associated with increased symptom severity compared to other transmission routes.

##### S2.2.3 Adenovirus

Adenoviruses (AdVs) are pathogens that can cause infection in the URT and LRT, in addition to the eyes and gastrointestinal tract. They are highly prevalent in children, with one US study finding AdVs accounted for 11% of 2638 lower respiratory tract infections in patients enrolled across three children's hospitals [100]. AdVs generally produce mild, "cold-like" symptoms but can potentially cause severe, or even fatal, disease in children and adults [101]. However, severe disease in immunocompetent adults is rare. Between 1971 and 2006, only 14 cases worldwide were recorded as being severe enough to result in respiratory failure [102]. Five of these cases were recorded in military barracks where clusters of severe cases have regularly been reported [103, 104] and six resulted from an outbreak in a mental health care facility [105]. These clusters of severe cases could be explained by symptom propagation.

**Dose-severity relationship**

We found evidence that those with more severe symptoms had a significantly higher AdV viral load in all identified studies (4/4) [106–109]. We found no studies exploring whether a higher viral load was associated with infecting others with a larger infectious dose. Whether infection with a larger infectious dose results in more severe symptoms remains an open question. Of the two studies identified, one [110] found a relationship, but the other [111] did not.

*More severe cases have a higher pathogen load*

Hospital cohort studies indicate that initial respiratory tract viral load is significantly correlated with both severity [108] and mortality [109]. In addition, Zecca *et al.* [106] found a significant correlation between peak viral load and mortality in a hospital cohort study of 241 AdV-infected patients; each $\log_{10}$ increase in viral load was associated with a 47% (95% CI 29%-67%) rise in the hazard for mortality. Goikhman *et al.* [107] also found a significant correlation between various severity scores and viral load in a study of 123 patients.

*Inconclusive evidence that infection with a larger dose results in more severe symptoms*

Teunis *et al.* [110] reviewed five AdV human challenge studies with a total of 91 volunteers and concluded that higher doses are necessary for acute symptoms to occur. In contrast, Couch *et al.* [111] performed a dose-varying human challenge study with 37 volunteers and found no relationship between severity and dose.

**Route-severity relationship**

There is support in the literature for AdV spread by both close contact [112, 113] and aerosol transmission [112, 114, 115]. We found no studies exploring whether more severe symptoms were associated with increased aerosol production. However, all identified studies (4/4) [111, 116–118] found that infection in the URT was generally less severe than infection by aerosols.

*Infection via aerosols is more likely to result in severe symptoms*

Human challenge studies give evidence for severity depending on the route of transmission. Couch *et al.* [111], inoculated 21 volunteers via small aerosols (1.5 $\mu$m) and 16 volunteers intranasally, using a range of inoculant doses. They found that intranasal inoculation required a 70-fold larger dose than aerosolised inoculation for infection to occur. Additionally, even among individuals who tested positive for AdV infection after intranasal inoculation, symptoms rarely developed; in contrast, almost all (15) of those inoculated via aerosols were symptomatic, and those symptoms were more severe. This evidence led them to suggest that infection of the lower respiratory tract is a requirement for severe symptoms and to point to aerosols produced by coughs and sneezes as a vector through which such infection could occur. This study is consistent with evidence from their earlier work [116], and with a review by Bell *et al.* [117] of 13 human challenge studies and a study involving intranasal inoculation of 13 volunteers [118] which found that intranasal inoculation tended to result in asymptomatic or mild upper respiratory illness, with severe symptoms rarely occurring.

#### S2.3 Viruses that cause pox-like illness

##### S2.3.1 Variola virus (Smallpox)

Smallpox is a disease caused by the variola virus. It is particularly known for the widespread use of 'variolation' to gain immunity. Variolation is where material from infected individuals (such as scabs) are either inhaled or rubbed into small cuts in the skin [119]. Variolation was occasionally fatal, but to a much lesser extent than naturally acquired smallpox [119]. The success of variolation and subsequent vaccine development allowed smallpox to be eradicated in 1980 [120]. Smallpox is the only human disease for which eradication has successfully been achieved.

###### Dose-severity relationship

We identified one study [121] exploring whether individuals with more severe symptoms had a viral load, which found a positive correlation. We found no studies exploring whether a higher viral load was associated with infecting others with a larger infectious dose. We identified one study [122] investigating the effect of a larger infectious dose on symptom severity, which found no such relationship.

*More severe cases have a higher pathogen load*

In a household-based study of 254 families, Rao *et al.* [121] found that URT viral load was the highest in those with the most severe symptoms.

*Inconclusive evidence that infection with a larger dose results in more severe symptoms*

Smallpox had a long latent period, with initial infection going undetected by the immune system [122]. This meant that, by the time the virus was detected, mass replication had already occurred, resulting in a high viral load regardless of the infectious dose [122].

###### Route-severity relationship

There has been limited discussion of variola transmission routes in the literature. However, in their review, Milton [123] give evidence for transmission through both aerosols and close contact. They also suggest that the severity of smallpox disease depends on the route of transmission, coining the term 'anisotropic infection' to mean when the transmission route determines either the virulence or probability of infection. Kiang and Krathwohl [124] also suggest the possibility of both aerosol and close contact transmission.

We found no studies exploring whether more severe symptoms was associated with increased aerosol production. However, all studies identified (4/4) [125–128] give evidence for infection via aerosols resulting in more severe symptoms that infection via other routes.

*Infection via aerosols is more likely to result in severe symptoms*

Hopkins [125] suggests that the site of infection determines the severity of symptoms, after finding that intranasal inoculation resulted in less severe symptoms than natural infection. This is supported by the finding that URT infection was associated with mild or asymptomatic infection; Sarkar *et al.* [126] found that only 4/34 individuals with positive URT samples developed symptomatic disease.

Animal model studies have found that aerosol transmission resulted in infection that was concentrated in the LRT [127] and was more severe compared to close contact transmission [128].

##### S2.3.2 Varicella zoster virus (Chickenpox)

Varicella zoster virus (VZV) is the causative agent of chickenpox (or varicella). VZV is highly contagious and around 77% of children in the UK are thought to be infected by age five [129]. Symptoms are notably more severe in older children and adults than in younger children [130]. After infection with VZV, the virus remains in the body as a latent infection [130]. This latent infection may be reactivated later in life causing herpes zoster (shingles), which can lead to serious complications [130].

###### Dose-severity relationship

We identified one study [131] which found that individuals with more severe symptoms had a higher viral load. However, we found no studies exploring whether a higher viral load was associated with infecting others with a larger infectious dose.

Anecdotally, there is a belief that within-household transmission of VZV results in more severe symptoms, due to an increased intensity of exposure and thus infectious dose [132]. Whilst there has been a limited exploration of this in the literature, we identified two studies [133] which found a significant correlation between intensity of exposure and symptom severity.

*More severe cases have a higher pathogen load*

Malavige *et al.* [131] found that blood viral load was significantly associated with a higher severity score ($r = 0.56, p = 0.0005$) in a study of 34 patients with confirmed VZV infections.

*Inconclusive evidence that infection with a larger dose results in more severe symptoms*

In a household-based study of 125 index cases and 40 secondary cases, Poulsen *et al.* [133] found that the mean number of pox (a measure of clinical severity) was significantly higher in secondary cases, compared to index cases ($p < 0.001$). However, they found no difference in fever response between index and secondary cases. Ross *et al.* [134] also found that secondary cases had a higher mean number of pox.

###### Route-severity relationship

Aerosols transmission has been found to contribute substantially to the spread of VZV [12, 135], with direct contact with infectious secretions being thought to be another potential route of transmission [136, 137].

We identified no studies exploring whether more severe symptoms were associated with increased aerosol production, or whether infection via aerosols was associated with increased symptom severity compared to other transmission routes.

#### S2.4 Bacteria

##### S2.4.1 *Mycobacterium tuberculosis*

*Mycobacterium tuberculosis* (Mtb) is the causative agent of Tuberculosis disease (TB), which in 2019 (before the COVID-19 pandemic) was the leading cause of death worldwide from an infectious disease, surpassing HIV/AIDS [138].

Typically, TB cases are categorised as either active or latent, where latent cases are asymptomatic and not infectious [139]. Minimal emphasis is typically put on asymptomatic cases and many policies are based on reported symptoms, meaning asymptomatic cases are generally overlooked [140]. However, there is growing evidence that a large proportion of infectious TB cases may never develop clinical symptoms. In their review, Frascella *et al.* [141] found that between 36.1% and 79.7% (median, 50.4%) of bacteriologically confirmed TB cases were subclinical (where subclinical was defined as asymptomatic). These infectious asymptomatic cases have only recently begun to be uncovered, and little is known about how their transmission differs from symptomatic cases.

###### Dose-severity relationships

We found evidence that individuals more severe symptoms had a higher bacterial load in all studies identified (8/8) [142–149], although, in two of these studies [143, 147] the findings were not all significant. In another [149] significance was not tested. All studies included (3/3) [148, 150, 151] found that those with a higher bacterial load also produced a significantly larger number of infectious aerosols. However, two of these studies [148, 150] found no positive correlation between aerosol production and symptom severity, with one [148] finding a negative correlation. We found no studies investigating the relationship between inoculant dose and severity, although two studies [150, 152] found a positive relationship between the inoculant dose and likelihood of developing active TB disease.

*More severe cases have a higher pathogen load*

Shimazaki *et al.* [142] found a significant and independent association between bacterial load (measured by PCR) and two-week mortality, and [150] found that a large bacterial load was significantly correlated with a number of severity markers, including length of illness ($p = 0.04$) and sputum volume ($p = 0.01$). Similarly, Du Bruyn *et al.* [144] found a significant correlation between Ct value and the Timika radiographic severity score [153] ($p < 0.004$). Bacterial load was also found to correlate with mortality in an animal model study involving Guinea pigs [149].

Many studies have also used time to positivity (TTP) as a proxy for bacterial load, as they are inversely correlated [154]. These studies show a significant inverse relationship between TTP and (i) the presence of lung cavities [145, 146], and (ii) the cavity score (a measure of the number and size of cavities) [147]. Theron *et al.* [148] also found a significant correlation between the presence of cavities and bacterial load, and Jones-López *et al.* [150] found a similar correlation in their hospital-based study of 85 patients, but it was not significant ($p = 0.65$). Murthy *et al.* [145] found a significant relationship between the extent of disease and TTP, although they suggested that this association was small since a 10-fold increase in TTP was required for an 11.4% decrease in the area affected. In a hospital-based study of 97 patients, te Riele *et al.* [147] found a non-significant relationship between TTP and disease extent ($p = 0.071$).

*A higher pathogen load results in infecting others with a larger dose*

We would expect that those with a higher bacterial load would be more likely to infect others with a larger inoculant dose. Indeed, Theron *et al.* [148] found a positive correlation between bacterial load and aerosol colony forming units (CFUs), and Jones-López *et al.* [150] found those in the high bacterial load group had a significantly higher median aerosol CFU count than those in the low bacterial load groups ($p = 0.04$). Acuña-Villaorduña *et al.* [151] also found that those with high bacterial load were significantly more likely to be aerosol positive (defined by the detection of any Mtb aerosol CFUs) ($p = 0.01$).

Theron *et al.* [148] also found a significant positive correlation between strength of cough and aerosol positivity but a negative correlation between symptom score and aerosol positivity (p=0.054). Similarly, Jones-López *et al.* [150] found that aerosol CFU count was correlated with strength of cough ($p = 0.008$) but was not associated with any measure of TB disease progression. Theron *et al.* [148] call for more research into transmission from those with mild symptoms and suggest finding strategies that target patients with cough over those with severe symptoms.

The negative correlation between aerosol production and severity may be explained by the significant correlation between lung function and aerosol positivity ($p = 0.014$) found by Theron *et al.* [148] as those with more severe symptoms may have reduced lung function. Alternatively, it may be that infection has spread outside of the lungs in those with more severe symptoms. One animal model study in guinea pigs found that, whilst bacterial load was correlated with mortality, the extent to which Mtb had spread outside of the lungs to other vital organs was a more important measure for predicting mortality [149].

*Infection with a larger dose results in more severe symptoms*

Jones-López *et al.* [150] found that those in contact with a high-aerosol case (an individual with a high aerosol CFU count) were significantly more likely to be infected and to develop TB disease than if they were in contact with a low-aerosol case. However, they did not explore the severity of symptoms in these secondary cases.

In their review, Fennelly and Jones-López [152] explore the relationship between inoculant dose and the development of latent TB. They find evidence that those infected with a lower inoculant dose are more likely to develop latent TB than if they had been infected with a higher dose. As latent cases are not infectious, it is unclear how this finding contributes to symptom severity, since those with latent infection cannot pass on latent infection to others. However, the demonstrated relationship between inoculant dose and subsequent immune response supports a dose-severity relationship within active cases.

##### Route-severity relationships

TB has historically been considered the “archetypical” example of a pathogen spread via aerosols and it has often been assumed that this is the only transmission route possible [155, 156]. However, it has been suggested that other transmission routes have been overlooked and may play a role in the transmission of Mtb [152]. Despite this suggestion, transmission routes seem unlikely to contribute to symptom propagation.

Whilst large droplet transmission can occur, it does so rarely, with one study finding that 96% of culturable Mtb was in particles smaller than $4.7\mu m$ [157]. In addition, Rohwedder [158] found that the URT was involved in less than 2% of TB cases, and this involvement is likely as a complication of LRT infection, as opposed to infection being initiated in the URT [158, 159]. However, it has been suggested that URT TB has historically been misdiagnosed and thus may play a larger part in TB infection than previously thought [160]. Indeed, Fennelly and Jones-López [152] suggest that infection of the URT by large droplets occurs relatively often and contributes to the widespread latent TB infection.

We found no studies exploring whether more severe symptoms were associated with increased aerosol production. We also found no studies directly investigating whether infection via aerosols was associated with increased symptom severity compared to infection via other routes. However, Fennelly and Jones-López [152] found that large droplet transmission was more likely to result in latent TB disease than aerosol transmission. Whilst latent cases are not infectious and thus cannot directly contribute to symptom propagation, this finding suggests that infection via large droplets is more likely to result in infection with a reduced immune response, which could result in infectious mild or asymptomatic cases.

##### S2.4.2 *Bordetella pertussis*

*Bordetella pertussis* are highly transmissible bacteria, being the causative agent of pertussis (also known as whooping cough) that has historically caused high mortality levels. *B. pertussis* primarily affects infants, with hospitalisations and deaths primarily being in infants [161]. The skew of health impacts towards infants has motivated widespread vaccination campaigns targeting infants six to eight weeks of age. Despite high levels of immunity in the population, there has been a rise in reported cases over the last two decades in high-income countries such as the USA [162], making it a pathogen of interest.

**Dose-severity relationship**

We found evidence that individuals with more severe disease had a significantly higher bacterial load in all studies identified (3/3) [163–165]. We found no studies exploring whether a higher bacterial load was associated with infecting others with a larger infectious dose. We also found no studies explicitly investigating the effect of a larger infectious dose on disease severity, but we found one study [166] which found a significant correlation between intensity of exposure and severity.

*More severe cases have a higher pathogen load*

Brotons *et al.* [163] found that median bacterial load was significantly higher in hospitalised patients ($n = 246$) than in non-hospitalised individuals ($n = 75$) and in those with complications (mostly classified as pneumonia) ($n = 9$). They also found a weak but significant correlation between median bacterial load and the number of symptoms on presentation ($r = 0.13, p = 0.002$). Similarly, Bolotin *et al.* [164] found hospitalised patients ($n = 22$) had a significantly higher bacterial load (indicated by low Ct value) than non-hospitalised individuals ($n = 202$, median Ct values of 20.7 vs. 31.6, $p < 0.001$). DeVincenzo *et al.* [165] found a significant correlation between bacterial load on diagnosis and length of hospital stay ($p = 0.0003$).

*Infection with a larger dose results in more severe disease*

Nielsen *et al.* [166] suggested that intensity of exposure was a risk factor for severe disease after finding significantly higher mortality in secondary cases caused by within-household transmission, compared to primary cases. This finding suggests a higher infecting dose may lead to more severe disease.

**Route-severity relationship**

There is evidence to support the claim that *B. pertussis* is transmitted via aerosol particles [61, 167–169]. However, there has been minimal exploration of other routes of transmission in the literature. Although it has previously been assumed that transmission via infected surfaces is impossible, [170], *B. pertussis* has been shown to survive on surfaces for between one to five days and on an individual's skin for up to six hours [171]. *B. pertussis* was also one of a number of pathogens detected during a pilot study investigating the possibility of mobile phones acting as pathogen reservoirs [172]. If contact transmission of *B. pertussis* is possible, this transmission route would likely play a larger role in asymptomatic transmission. For symptomatic cases, the characteristic "whooping cough" may aid in aerosol transmission.

We found no studies exploring whether more severe disease was associated with increased aerosol production. However, we found two studies [173, 174] that found more severe disease after LRT infection, compared to URT infection.

*Infection via aerosols is more likely to result in severe disease*

Animal model studies give evidence for a relationship between the site of inoculation and the severity of disease. Soumana *et al.* [173] found that inoculation of the lower respiratory tract (LRT) in mice led to severe disease with infection remaining localised in the lungs. In contrast, when the mice were inoculated in the upper respiratory tract (URT), infection was much milder and more prolonged with bacterial particles rarely reaching the lungs. Another study in juvenile baboons found that inoculation in the LRT led to severe disease with pathology similar to that seen in fatal human infant cases [174].

##### S2.4.3 Group A streptococci (GAS)

Group A streptococci (GAS) is a collection of bacterial pathogens. Although multiple species of streptococcus may fall into this group, GAS is usually used to refer to *Streptococcus pyogenes* specifically. GAS can cause a wide range of clinical manifestations. Typically, GAS infections are mild, URT infections [175]. However, more severe manifestations can also occur in the case of invasive GAS (iGAS) infections. iGAS has historically been associated with more systemic severe symptoms such as toxic shock syndrome or necrotising fasciitis (also known as "flesh-eating disease") [175]; recently, however, there has been an increase in the proportion of iGAS cases associated with LRT infection and pneumonia [176, 177].

The factors surrounding how iGAS prevents clinically are not fully understood, although it is thought to be highly dependent on host genetic factors [178]. There are also notable variations in pathogenicity between strains [177]; one particular pathogenic strain is thought to be the reason for a cluster of severe iGAS cases within a household [179].

###### Dose-severity relationships

We identified three studies [180–182] exploring whether those with more severe disease have higher bacterial loads of which two found a positive association [181, 182]. We found no studies exploring whether a higher bacterial load was associated with infecting others with a larger infectious dose. Similarly, we found no studies investigating the effect of a larger infectious dose on symptom severity.

*Inconclusive evidence that more severe cases have a higher pathogen load*

In a hospital-based study of 24 patients, Dunne *et al.* [180] found no association between URT bacterial load and severity score. However, in biopsy studies, both Thulin *et al.* [181] and Norrby-Teglund *et al.* [182] found that bacterial load was correlated with tissue inflammation.

###### Route-severity relationships

GAS is generally thought to be transmitted through large droplets and direct contact transmission [175, 183]. However, it has been suggested that transmission via aerosols may contribute to GAS transmission [184]. Whilst aerosol transmission has not yet been confirmed, a human challenge study is currently being undertaken to investigate its relative contribution [185].

Whilst initial URT infection is known to be common for GAS, and often more severe LRT symptoms are thought to occur from LRT invasion post-URT infection, the potential for direct infection of the LRT has not been explore and cannot be excluded.

We found whether more severe disease was associated with increased aerosol production. We also found no studies directly investigating whether infection via aerosols was associated with increased disease severity compared to other transmission routes. However, animal model studies have found that LRT infection, either through aerosol or direct LRT inoculation, led to severe LRT infection with pathology similar to that seen in human iGAS cases [186, 187].
