## Supplementary Material S2 for "Symptom propagation in respiratory pathogens of public health concern: a review of the evidence"

### Supplementary File S2: Summary Tables

#### Symptom propagation in respiratory pathogens of public health concern: a review of the evidence

Phoebe Asplin 1,2,3*, Rebecca Mancy 4,5, Thomas Finnie 6, Fergus Cumming 7, Matt J. Keeling 2,3,8, Edward M. Hill 2,3*

1 EPSRC & MRC Centre for Doctoral Training in Mathematics for Real-World Systems, University of Warwick, Coventry, Unit
2 Mathematics Institute, University of Warwick, Coventry, United Kingdom.
3 The Zeeman Institute for Systems Biology & Infectious Disease Epidemiology Research, University of Warwick, Coventry, U
4 School of Biodiversity, One Health and Veterinary Medicine, University of Glasgow, Glasgow, United Kingdom.
5 MRC/CSO Social and Public Health Sciences Unit, University of Glasgow, Glasgow, United Kingdom.
6 Data, Analytics and Surveillance, UK Health Security Agency, London, United Kingdom.
7 Foreign, Commonwealth and Development Office, London, United Kingdom.
8 School of Life Sciences, University of Warwick, Coventry, United Kingdom.

This file contains the summary tables for all of the studies included in the review 'Symptom propagation in respiratory pathogens of public health concern: a review of the evidence'. Studies are grouped by pathogen and then by whether they correspond to dose-severity or route severity relationships. Studies are then classified as either for or against the hypothesis, with mixed studies containing findings that are both for and against. They are also then given a strength of evidence classification: high - a study directly investigating symptom propagation with significant findings; moderate - a study strongly related to part of the mechanism with significant findings; low - a study with either non-significant findings, or that is more weakly related to part of the mechanism.

**Table of Contents**

S1 Pathogen non-specific 2
S2 Adenovirus 3
S3 *Bordetella pertussis* 4
S4 Group A streptococci 5
S5 Influenza virus 6
S6 Measles virus 10
S7 MERS-CoV 12
S8 *Mycobacterium tuberculosis* 14
S9 Rhinovirus 16
S10 Respiratory syncytial virus 18
S11 SARS-CoV-1 21
S12 SARS-CoV-2 22
S13 Variola virus 26
S14 Variola zoster virus 27
S15 *Yersinia pestis* 28

### S1 Pathogen non-specific

| Dose-severity relationship | | | | | |
|---|---|---|---|---|---|
| **Paper** | **Study Type** | **Study size** | **Key findings** | **For/against** | **Strength of evidence** |

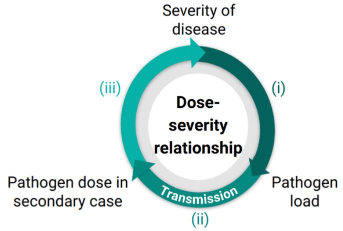

| Route-severity relationship | | | | | | |
|---|---|---|---|---|---|---|
| **Paper** | **Study Type** | **Study size** | **Key findings** | **For/against** | **Strength of evidence** | |
| Fabian et al (2011). "Origin of Exhaled Breath Particles from Healthy and Human Rhinovirus-Infected Subjects" | Community based | 19 | Exhaled particle concentrations peaked at the end of an exhalation, suggesting that they are generated by the reponening of collapsed small airways or alveoli. Exhaled particle concentrations were significantly correlated with minute ventilation (the volume of air that enters the lung in a minute) but was not correlated with breath frequency (r=0.26). These findings suggest that breath aerosols originate from the LRT. They also found that exhaled particle concentrations correlated with airflow during the first half of exhalation, suggesting that some exhaled droplets originate from the URT. Smaller droplets, i.e. aerosols, (0.3-0.499 mu m) correlated with airflow and continued increasing towards the end of exhalation; larger droplets (0.5-1 mu m) had a better correlation with airflow at the start of exhalation and increased only slightly at the end of exhalation. This suggests that aerosols primarily orginiate from the LRT, where as large droplets primarily originate from the URT | For | Moderate | i) Aerosols primarily originate from the LRT |
| Almstrand et al (2010). "Effect of airway opening on production of exhaled particles" | Community based | 10 | Deep exhalation resulted in the production of a much larger number of particles than shallow breathing. Additionally, when exhalation was halted at the closing point (the point at which airways begin to close during exhalation), substantially fewer particles were produced, despite a minimal decrease in the volume of air exhaled. These findings suggest that exhaled particles are produced during airway closure which occurs in the terminal bronchioles (in the LRT) | For | Moderate | |
| Johnson et al (2011). "Modality of human expired aerosol size distributions" | Community based | 15 | Droplets produced during speech and cough have three distinct droplet size distribution modes; 1.3, 2.0 and 145 mu m for speech and 1.3, 1.4 and 123 mu m for cough. These three modes are associated with distinct processes that occur in different parts of the respiratory tract. Namely, the deep respiratory tract, the larynx and the URT. | For | Moderate | |
| Johnson & Morawaska (2009). "The Mechanism of Breath Aerosol Formation" | | | Deep exhalation resulted in a 5-fold increase in the average concentration of particles. Increasing the exhalation rate had no effect. These findings support the hypothesis that small particles are generated in the LRT, rather than being generated by turbulence in the airways | For | Moderate | |
| Morawaska et al (2009). "Size distribution and sites of origin of droplets expelled during expiratory activities" | Community based | 15 | Coughing and speech produce large droplets from vibrations of vocal cords | For | Low | |
| Hatch (1961). "Distribution and deposition of inhaled particles in respiratory tract" | Particle mechanics | n/a | Close to 100% of droplets larger than 10 mu m and 80% of 5 mu m droplets will deposit in the URT. This proportion approaches zero when droplets are sized between 1-2 mu m | For | Moderate | ii) Infection via aerosols result in LRT infection |
| Licht (1972). "The Movement of Aerosol Particles" | Particle mechanics | n/a | Most droplets larger than 2 mu will not reach the LRT whereas particles less tha 1 mu essentially never deposit in the URT. | For | Moderate | |
| Lippman & Albert (2007). "The Effect of Particle Size on the Regional Deposition of Inhaled Aerosols in the Human Respiratory Tract" | Human challenge | 34 | URT and tracheobronchial deposition increased with particle size, whereas alveolar deposition decreased. | For | Moderate | |
| Emmett et al (1982). "Measurements of the total and regional deposition of inhaled particles in the human respiratory tract" | Human challenge | 12 | Deposition in the alveoli was higher for 3.5 mu particles, compared to 5.2 mu, 8 mu and 10 mu particles, despite the total deposition across the respiratory tract being decreased | For | Moderate | |

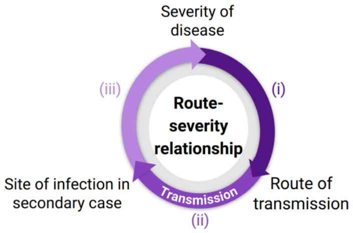

### S2 Adenovirus

**Dose-severity relationship**

| Paper | Study Type | Study size | Key findings | For/against | Strength of evidence | |
|---|---|---|---|---|---|---|
| Zecca et al (2019). "Association between adenovirus viral load and mortality in pediatric allo-HCT recipients: the multinational AdVance study" | Hospital based | 241 | Viral load in stem cell transplant recipients significantly correlated with all-cause mortality (OR 1.47; 95% CI 1.29-1.67 per log10 increase in viral load). | For | Moderate | i) More severe cases have a higher pathogen load |
| Goikhman et al (2020). "Adenovirus load correlates with respiratory disease severity among hospitalized pediatric patients" | Hospital based | 123 | Ct value significantly, negatively correlated with severity score | For | Moderate | |
| Xie et al (2018). "Human adenovirus load in respiratory tract secretions are predictors for disease severity in children with human adenovirus pneumonia" | Hospital based | 174 | Viral load significantly correlated with severity of pnuemonia (r = 0.477, p = 0.000) | For | Moderate | |
| Gu et al (2016). "Sustained Viremia and High Viral Load in Respiratory Tract Secretions Are Predictors for Death in Immunocompetent Adults with Adenovirus Pneumonia" | Hospital based | 14 | Initial viral load significantly correlated with mortality | For | Moderate | |
| Teunis et al (2016). "A generalized dose-response relationship for adenovirus infection and illness by exposure pathway" | Review of human challenge studies | 91 across 5 studies | Larger doses are a requirement for severe symptoms to occur. Note that the route of inoculation varies between studies, with both aerosol and intranasal inoculation being used. | For | Low | iii) Infection with a larger dose results in more severe disease |
| Couch et al (1969). "The minimal infectious dose of adenovirus type 4; the case for natural transmission by viral aerosol" | Human challenge | 37 | No relationship found between inoculant dose and severity after intranasal or aerosol inoculation | Against | Low | |

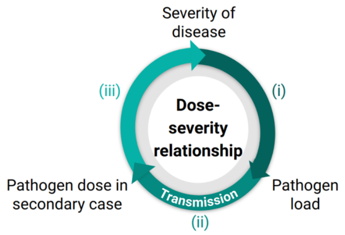

**Route-severity relationship**

| Paper | Study Type | Study size | Key findings | For/against | Strength of evidence | |
|---|---|---|---|---|---|---|
| Couch et al (1969). "The minimal infectious dose of adenovirus type 4; the case for natural transmission by viral aerosol" | Human challenge | 37 | Aerosol inoculation required a 70-fold lower dose than intranasal inoculation for infection to occur and resulted in symptomatic disease more often and more severe symptoms | For | Moderate | iii) Infection in the LRT (i.e. via aerosols) results in more severe disease than URT infection |
| Couch et al (1966). "Effect of route of inoculation on experimental respiratory viral disease in volunteers and evidence for airborne transmission" | Human challenge | 15 | Aerosol inoculation resulted in severe disease more often than large droplet inoculation (6/9 vs 3/6) | For | Low | |
| Bell et al (1956). "Studies of Adenoviruses (APC) in Volunteers" | Review of human challenge studies | 257 | Intranasal inoculation tended to result in mild or asymptomatic infection | For | Low | |

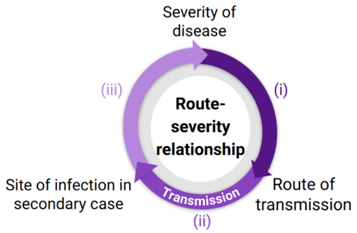

### S3 *Bordetella pertussis*

**Dose-severity relationship**

| Paper | Study Type | Study size | Key findings | For/against | Strength of evidence | |
|---|---|---|---|---|---|---|
| Brotons et al (2016). "Differences in Bordetella pertussis DNA load according to clinical and epidemiological characteristics of patients with whooping cough" | Hospital based | 321 | Bacterial load significantly correlated with hospitalisation and complications (mostly classified as pnuemonia). Bacterial load also weakly correlated with number of symptoms on presentation | For | Moderate | **i) More severe cases have a higher pathogen load** |
| Bolotin et al (2015). "Correlation of Real Time PCR Cycle Threshold Cut-Off with Bordetella pertussis Clinical Severity" | Hospital based | 627 | Ct value significantly correlated with hospitalisation, but not the presence of certain symptoms | For | Moderate | |
| DeVincenzo et al (2013). "Molecular detection and quantification of pertussis and correlation with clinical outcomes in children" | Hospital based | 104 | Ct value significantly correlated with length of hospital stay | For | Moderate | |
| Nielsen et al (2001). "Intensity of Exposure and Severity of Whooping Cough" | Hospital based | 269 | Secondary cases caused by within-household transmission had significantly higher mortality, compared to primary cases | For | Moderate | **iii) Infection with a larger dose results in more severe disease** |

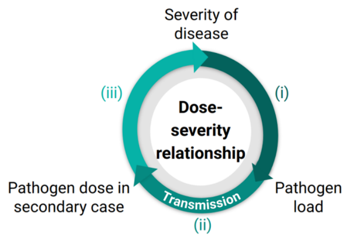

**Route-severity relationship**

| Paper | Study Type | Study size | Key findings | For/against | Strength of evidence | |
|---|---|---|---|---|---|---|
| Soumana et al (2021). "Modeling Immune Evasion and Vaccine Limitations by Targeted Nasopharyngeal Bordetella pertussis Inoculation in Mice" | Animal model (Mice) | 12 | LRT inoculation led to severe disease that remained localised in the lungs whereas after URT inoculation, the symptoms were milder and bacteria rarely reached the lungs | For | Low | **iii) Infection in the LRT (i.e. via aerosols) results in more severe disease than URT infection** |
| Zimmerman et al (2018). "Histopathology of Bordetella pertussis in the Baboon Model" | Animal model (Baboon) | Not stated | LRT inoculation led to severe disease with pathology similar to that seen in fatal human infant cases | For | Low | |

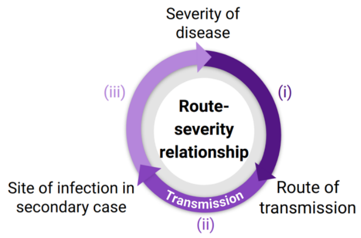

### S4 Group A streptococci

| Dose-severity relationship | | | | | |
|---|---|---|---|---|---|
| **Paper** | **Study Type** | **Study size** | **Key findings** | **For/against** | **Strength of evidence** |
| Dunne et al (2013). "Detection of group a streptococcal pharyngitis by quantitative PCR" | Hospital based | 24 | Found no association between pharyngeal bacterial load and severity score | Against | Low |
| Thulin et al (2006). "Viable group A streptococci in macrophages during acute soft tissue infections" | Biopsy | 70 | Highest levels of bacterial load significantly correlated with tissue inflammation (p<0.01) | For | Low |
| Norby-Teglund et al (2001). "Evidence for Superantigen Involvement in Severe Group A Streptococcal Tissue Infections" | Biopsy | 31 | Bacterial load correlated with the severity of tissue infection | For | Low |

**i) More severe cases have a higher pathogen load**

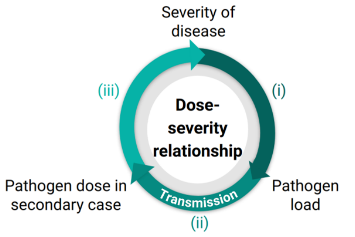

| Route-severity relationship | | | | | |
|---|---|---|---|---|---|
| **Paper** | **Study Type** | **Study size** | **Key findings** | **For/against** | **Strength of evidence** |
| Olsen et al (2010). "Lower respiratory tract infection in cynomolgus macaques (Macaca fascicularis) infected with group A Streptococcus" | Animal model (cynomolgus macaques) | 6 | Direct inoculation of the lungs resulted in severe pneumonia, with similar pathology to that seen in human cases. | For | Low |
| Hokonohara et al (1988). "Experimental studies on the initial focus of invasion of group A streptococci" | Animal model (rabbits) | Not stated | Aerosol inoculation resulted in LRT infection that closely resembled the pathoglogy in human invasive GAS cases. | For | Low |

**iii) Infection in the LRT (i.e. via aerosols) results in more severe disease than URT infection**

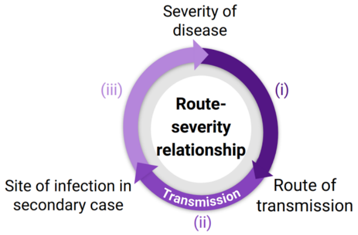

### S5 Influenza virus

| Dose-severity relationship | | | | | | |
|---|---|---|---|---|---|---|
| **Paper** | **Study Type** | **Study size** | **Key findings** | **For/against** | **Strength of evidence** | |
| Hall et al (1979). "Viral Shedding Patterns of Children with Influenza B Infection" | Hospital based | 43 | Peak influenza B URT viral load was significantly correlated with symptom severity and fever score | For | Moderate | **i) More severe cases have a higher pathogen load** |
| Hijano et al (2019). "Clinical correlation of influenza and respiratory syncytial virus load measured by digital PCR" | Hospital based | 23 | URT viral load was significantly correlated with fever (only significant for influenza A, not B), cough and nasal congestion | For | Moderate | |
| Lee et al (2009). "Viral Loads and Duration of Viral Shedding in Adult Patients Hospitalized with Influenza" | Hospital based | 147 | URT viral load on presentation was significantly higher in hospitalised patients, compared to time-matched outpatients | For | Moderate | |
| Lee et al (2013). "Influenza virus load in hospitalised patients" | Hospital based | 128 | URT viral load was significantly correlated with symptom score (r=0.219, p=0.01) and hospitalisation (p=0.003) | For | Moderate | |
| Fuller et al (2013). "Association of the CT values of real-time PCR of viral upper respiratory tract infection with clinical severity, Kenya" | Hospital based | 550 | HIgh URT Ct value (low viral load) was significantly associated with being an outpatient (OR 1.07, 95% CI 1.01-1.13) | For | Moderate | |
| Granados et al (2017). "Influenza and rhinovirus viral load and disease severity in upper respiratory tract infections" | Hospital based | 561 | Patients in the hospital/ICU had signficantly higher mean URT viral loads than patients in ambulatory settings for Influenza B (OR 1.28, 95% CI 1.11–1.47) and Influenza A (OR 1.48, 95% CI 1.25–1.75) | For | Moderate | |
| Yan et al (2018). "Infectious virus in exhaled breath of symptomatic seasonal influenza cases from a college community" | Community based | 142 | URT viral load was significantly associated with both URT and LRT symptoms | For | Moderate | |
| Launes et al (2012). "Viral load at diagnosis and influenza A H1N1 (2009) disease severity in children" | Hospital based | 93 | Ct value from URT samples on diagnosis significantly negatively correlated with requirement of mechanical ventilation | For | Moderate | |
| Lalueza et al (2019). "Influence of viral load in the outcome of hospitalized patients with influenza virus infection" | Hospital based | 239 | Ct value from URT samples significantly negatively correlated with abnormal findings on chest X-ray but not worse prognosis | For | Low | |
| Pronier et al (2021). "Respiratory Influenza viral load as a marker of poor prognosis in patients with severe symptoms" | Hospital based | 118 | Ct value from URT samples significantly negatively correlated with mortality | For | Moderate | |
| Couch et al (1971). "Correlated Studies of a Recombinant Influenza-Virus Vaccine. III. Protection against Experimental Influenza in Man" | Human challenge | 18 | Mean URT viral load significantly correlated with symptom severity (r=0.878, p<0.001) | For | Moderate | |
| Boon et al (2011). "H5N1 Influenza Virus Pathogenesis in Genetically Diverse Mice Is Mediated at the Level of Viral Load" | Animal model (mice) | 21 | Lung viral load significantly correlated with markers of inflammation | For | Moderate | |

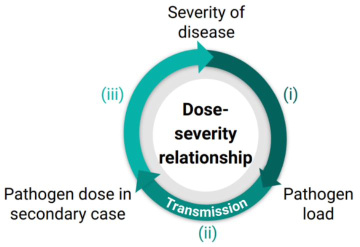

| | | | | | | |
|---|---|---|---|---|---|---|
| Spencer et al (2016). "Factors associated with real-time RT-PCR cycle threshold values among medically attended influenza episodes" | Community based | 2,466 | High URT viral load (indicated by Ct value <=23) was significantly associated with self-rated illness severity for influenza A. Individuals were significantly more likely to have a high viral load if they had very severe (OR 2.63; 95% CI 1.53-4.52), severe (OR 2.46; 95% CI 1.56-3.88) or moderate (OR 2.44; 95% CI 1.54-3.86) disease compared to those with mild disease. High viral load was also significantly associated with the presence of fever for influenza A (OR 1.89; 95% CI 1.28-2.78), but the association was not significant for influenza B (OR 1.67; 95% CI 0.95-2.92). Similarly, the association between high viral load and self-rated severity for influenza B was only significant for severe disease (OR 1.92; 95% CI 1.07-3.45), not for very severe disease (OR 1.21; 95% CI 0.59-2.48) or moderate disease (OR 1.48; 95% CI 0.82-2.68). | For | Moderate | |
| Rodrigues Guimarães Alves et al (2020). "Influenza A(H1N1)pdm09 infection and viral load analysis in patients with different clinical presentations" | Hospital based | 162 | URT Influenza A viral load on enrollment significantly higher in symptomatic patients than asymptomatics but there was no signficant difference between symptomatic outpatients and hospitalised patients | Mixed | | |
| To et al (2010). "Delayed Clearance of Viral Load and Marked Cytokine Activation in Severe Cases of Pandemic H1N1 2009 Influenza Virus Infection" | Hospital based | 74 | Individuals with severe disease (defined by the development of acute respiratory distress syndrome) have a slower decline in viral load. However, initial URT viral load was higher in fatal cases than in severe non-fatal cases or mild cases (5.45 (2.65-7.49), 5.68 (2.65–8.65), 5.97 (2.65–10.7) log10 copies/mL respectively). Note, for the ARDS groups, samples were taken on average 4 days after symptom onset vs 2 days after symptom onset for the mild cases. Tracheal viral load was also lower in fatal cases than severe but not fatal cases (4.97 (2.65–7.16) vs 5.24 (4.83–5.86) log10 copies/mL, respectively) | Against | Low | |
| Yan et al (2018). "Infectious virus in exhaled breath of symptomatic seasonal influenza cases from a college community" | Community based | 142 | URT viral load was not a significant predictor of viral load in either large droplets (p=0.48) or aerosols (p=0.16) | Against | Low | **ii) A higher pathogen load results in infecting others with a larger dose** |
| Koster et al (2012). "Exhaled Aerosol Transmission of Pandemic and Seasonal H1N1 Influenza Viruses in the Ferret" | Animal model (ferrets) | Not stated | Ferrets with higher URT viral loads were more transmissible (no transmission detected (0/3) from low viral load donor but transmission detected in all recipients (3/3) from a high viral load donor). However, viral load in exhaled aerosols was not found to be associated with transmissibility despite aerosol transmission being the only possible transmission route | For | Low | |
| Marois et al (2012). "Initial infectious dose dictates the innate, adaptive, and memory responses to influenza in the respiratory tract" | Animal model (mice) | Not stated | Relationship between intranasal inoculant dose and severity | For | Low | |
| Miller et al (2013). "The virus inoculum volume influences outcome of influenza A infection in mice" | Animal model (mice) | Not stated | Relationship between intranasal inoculant dose and severity | For | Low | |
| Pinto et al (1969). "Pathogenesis of and recovery from respiratory syncytial and influenza infections in ferrets" | Animal model (ferrets) | Not stated | Relationship between intranasal inoculant dose and severity | For | Low | |
| Han et al (2019). "A Dose-finding Study of a Wild-type Influenza A(H3N2) Virus in a Healthy Volunteer Human Challenge Model" | Human challenge | 37 | Relationship between intranasal inoculant dose and symptom scores | For | Moderate | |
| Memoli et al (2015). "Validation of the Wild-type Influenza A Human Challenge Model H1N1pdMIST: An A(H1N1)pdm09 Dose-Finding Investigational New Drug Study" | Human challenge | 46 | Relationship between intranasal inoculant dose and symptom scores | For | Moderate | **iii) Infection with a larger dose results in more severe disease** |

| | | | | | |
|---|---|---|---|---|---|
| Handel et al (2018). "Exploring the impact of inoculum dose on host immunity and morbidity to inform model-based vaccine design" | Within-host infection model | n/a | Morbidity monotonically increased with inoculant dose | For | Low |
| Chang et al (2007). "Simple scaling laws for influenza A rise time, duration, and severity" | Within-host infection model | n/a | Morbidity monotonically increased with inoculant dose above a threshold value of initial viral load (below the threshold value severity is constant with respect to initial viral load) | For | Low |
| Hancioglu et al (2007). "A dynamical model of human immune response to influenza A virus infection" | Within-host infection model | n/a | Morbidity monotonically increased with inoculant dose above a threshold value of initial viral load (below the threshold value severity is constant with respect to initial viral load) | For | Low |
| Carrat et al (2008). "Time Lines of Infection and Disease in Human Influenza: A Review of Volunteer Challenge Studies" | Meta-analysis | 56 studies, 1280 participants | Found a non-significant association between inoculant dose and symptomatic infection (p=0.12) and a significant association between inoculant dose and presence of fever (OR = 0.56, 95%CI: 0.42-0.73) | Against | Low |

#### Route-severity relationship

| Paper | Study Type | Study size | Key findings | For/against | Strength of evidence | |
|---|---|---|---|---|---|---|
| Lindsley et al (2016). "Viable influenza A virus in airborne particles expelled during coughs versus exhalations" | Community based | 53 | Viable influenza was found more often in cough aerosols than breath aerosols, but the difference was not significant (p=0.2207) | For | Low | **i) More severe disease results in increased aerosol production** |
| Bischoff et al (2013). "Exposure to Influenza Virus Aerosols During Routine Patient Care" | Hospital based | 61 | Patients who reported severe symptoms produced significantly more influenza aerosols. Coughing and sneezing was associated with increased aerosol production, but only in those with a high viral load | For | Moderate | |
| Yan et al (2018). "Infectious virus in exhaled breath of symptomatic seasonal influenza cases from a college community" | Community based | 142 | Viral RNA in aerosols was significantly correlated with cough frequency. Viral load was associated with URT symptoms. Suggest that aerosols reflect infection in the LRT, since there was no significant association between nasalpharangeal viral load and aerosol production (p=0.16) | For | Moderate | |
| Bodewes et al (2011). "Pathogenesis of Influenza A/H5N1 Virus Infection in Ferrets Differs between Intranasal and Intratracheal Routes of Inoculation" | Animal model (Ferrets) | 8 | Aerosol inoculation resulted in LRT infection more often than intranasal inoculation | For | Low | **ii) Infection via aerosols is more likely to result in LRT infection** |
| Smith et al (2011). "Aerosol Inoculation with a Sub-lethal Influenza Virus Leads to Exacerbated Morbidity and Pulmonary Disease Pathogenesis" | Animal model (Mice) | Not stated | Aerosol inoculation resulted in more severe LRT infection than intranasal inoculation | For | Low | **iii) Infection in the LRT (i.e. via aerosols) results in more severe disease than URT infection** |
| Yetter et al (1980). "Outcome of Influenza Infection: Effect of Site of Initial Infection and Heterotypic Immunity" | Animal model (Mice) | 32 | Aerosol inoculation resulted in more severe LRT infection than intranasal inoculation | For | Low | |
| Mooji et al (2021). "Aerosolized Exposure to H5N1 Influenza Virus Causes Less Severe Disease Than Infection via Combined Intrabronchial, Oral, and Nasal Inoculation in Cynomolgus Macaques" | Animal model (Cynomolgus macaques) | 12 | Inoculation directly into the lungs produced more severe disease when compared to other studies that inoculated the trachea | For | Low | |
| Cowling et al (2013). "Aerosol transmission is an important mode of influenza A virus spread" | Household based | 782 | The risk of fever and cough when infected via the aerosol route was around twice that versus being infected via the contact route (53% vs 23% vs 22% for aerosol, droplet and direct contact respectively in Hong Kong households, 77% vs 33% vs 26% in Bangkok households) | For | Moderate | |

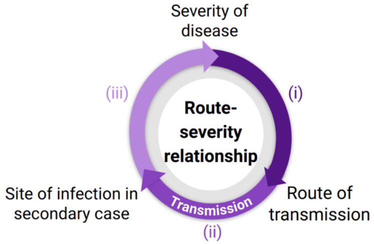

| | | | | | |
|---|---|---|---|---|---|
| Carrat et al (2008). "Time Lines of Infection and Disease in Human Influenza: A Review of Volunteer Challenge Studies" | Meta-analysis | 1,280 | Intranasal inoculation resulted in mild disease. | For | Low |
| Alford et al (1966). "Human Influenza Resulting from Aerosol Inhalation" | Human challenge | 11 | Aerosol inoculation resutled in a wide range of clinical symptoms, more similar to those seen in natural infections. In contrast, URT inoculation only produces mild infection with no LRT involvement | For | Low |

### S6 Measles virus

| Dose-severity relationship | | | | | | |
|---|---|---|---|---|---|---|
| **Paper** | **Study Type** | **Study size** | **Key findings** | **For/against** | **Strength of evidence** | |
| Aaby (1992). "Patterns of exposure and severity of measles infection copenhagen 1915–1925" | Hospital based | 221 | Individuals infected by someone with severe measles were significantly more likely to develop severe disease (OR 2.90; 95% CI 1.63-5.17) and their disease was more likely to be fatal (OR 3.87; 95% CI 1.65-9.08). Individuals exposed to two or more index cases also had a significantly higher CFR than those exposed to only one index case (OR 1.90; 95% CI 1.12-3.22) | For | High | **General** |
| Aaby & Leeuwenburg (1990). "Patterns of Transmission and Severity of Measles Infection: A Reanalysis of Data from the Machakos Area, Kenya" | Household based | 1037 | Significantly higher CFR in cases infected by a fatal index case (OR 4.69; 95% CI 1.64-13.41) and in individuals infected within the household compared to index cases (OR 3.00; 95\% CI 1.55-5.80). Individuals exposed to two or more index cases also had a higher CFR than those exposed to only one index case, but the difference was not significant (OR 2.47; 95% CI 0.93-6.56) | For | High | |
| Samb (1994). "Impact épidémiologique et démographique de la rougeole et de la vaccination contre la rougeole dans une zone rurale du Sénégal (Niakhar)" | Community based | Not stated | Infection by an individual with respiratory complications was significnatly more likely to result in respiratory complications (OR 1.74; 95% CI 1.16-2.60). Vaccinated cases produced less severe disease in those they infected, but the difference was not significant (OR 0.30; 95% 0.04-2.09). Severity was associated with the number of index cases but it was not signficant (OR 1.26; 95% CI 0.62-2.57). Fatal cases were more likely to cause fatal disease in those they infect, again not significant (OR 1.44; 95% CI 0.73-2.81) | For | High | |
| Sundell et al (2019). "Measles outbreak in Gothenburg urban area, Sweden, 2017 to 2018: low viral load in breakthrough infections" | Community based | 28 | Breakthrough infections (infections in those who have been vaccinated) have been shown to have a lower viral load and less severe symptoms than infected individuals who were not vaccinated | For | Low | **i) More severe cases have a higher pathogen load** |
| Garenne & Aaby (1990). "Pattern of Exposure and Measles Mortality in Senegal" | Household based | 1,500 | CFR increased exponentially with generations of infection and is significantly higher in individuals infected with the household compared to index cases | For | Moderate | **iii) Infection with a larger dose results in more severe disease** |
| Aaby et al (1986). "High measles mortality in infancy related to intensity of exposure" | Household based | 721 | CFR significantly higher in individuals infected with the household compared to index cases | For | Moderate | |
| Aaby (1988). "Severe Measles in Copenhagen, 1915–1925" | Hospital based | 2,208 | CFR significantly higher in individuals infected with the household compared to index cases | For | Moderate | |
| Koster (1988). "Mortality Among Primary and Secondary Cases of Measles in Bangladesh" | Household based | 160 | CFR significantly higher in individuals infected with the household compared to index cases | For | Moderate | |
| Aaby et al (1984). "Determinants of Measles Mortality in a Rural Area of Guinea-Bissau: Crowding, Age, and Malnutrition" | Household based | 163 | CFR was significantly higher in clustered cases (multiple infected within the same household) | For | Moderate | |
| Aaby et al (1984). "Overcrowding and intensive exposure as determinants of measles mortality" | Household based | 387 | CFR was significantly higher in clustered cases (multiple infected within the same household) | For | Moderate | |
| Lamb (1988). "Epidemic Measles in a Highly Immunized Rural West African (Gambian) Village" | Household based | 54 | CFR was significantly higher in clustered cases (multiple infected within the same household) | For | Moderate | |
| Aaby (1995). "Assumptions and contradictions in measles and measles immunization research: Is measles good for something?" | Review | n/a | CFR was significantly higher in clustered cases (multiple infected within the same household) in three studies | For | Moderate | |
| Aaby et al (1986). "Severe Measles in Sunderland, 1885: A European—African Comparison of Causes of Severe Infection" | Household based | 311 | CFR was higher in clustered cases (multiple infected within the same household) in three studies, but the difference was not significant (p=0.11) | For | Low | |
| Wear et al (1968). "Virus replication and ultrastructural changes after induction of encephalitis in mice by measles virus" | Animal model (Mice) | 134 | A larger inoculant dose resulted in higher mortality | For | Low | |

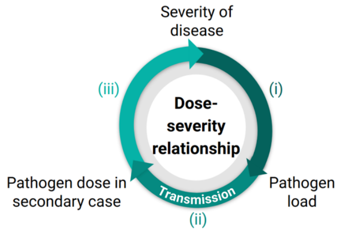

| Route-severity relationship | | | | | | |
|---|---|---|---|---|---|---|
| **Paper** | **Study Type** | **Study size** | **Key findings** | **For/against** | **Strength of evidence** | |
| de Vries et al (2012). "The pathogenesis of measles" | Review | n/a | URT cells are not the initial target of measles virus as these cells do not express CD150, which has been found to be a requirement for measles infection | Against | Moderate | **Infection is possible in both the LRT and URT** |
| Lemon et al (2011). "Early Target Cells of Measles Virus after Aerosol Infection of Non-Human Primates" | Animal model (Macaques) | 12 | Infection in the URT was not detected after aerosol inoculation until after infection had reached the bloodstream | Against | Moderate | |
| Ludlow et al (2013). "Measles Virus Infection of Epithelial Cells in the Macaque Upper Respiratory Tract Is Mediated by Subepithelial Immune Cells" | Animal model (Macaques) | 40 | Infection in the URT was not detected after aerosol inoculation until after infection had reached the bloodstream | Against | Moderate | |

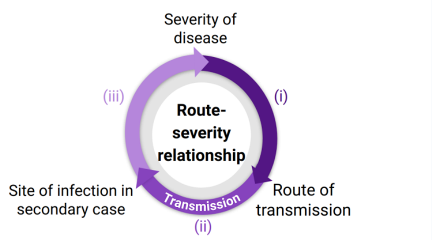

#### S7 MERS-CoV

| Dose-severity relationship | | | | | | |
|---|---|---|---|---|---|---|
| **Paper** | **Study Type** | **Study size** | **Key findings** | **For/against** | **Strength of evidence** | |
| Oh et al (2016). "Viral Load Kinetics of MERS Coronavirus Infection" | Hospital based | 17 | Viral load higher in those with severe disease. The relationship was significant for LRT samples (p<0.001), but not URT samples (p=0.06) | For | Moderate | **i) More severe cases have a higher pathogen load** |
| Feikin et al (2015). "Association of Higher MERS-CoV Virus Load with Severe Disease and Death, Saudi Arabia, 2014" | Hospital based | 102 | Ct value was a significant predictor of mortality | For | Moderate | |
| Min et al (2016). "Comparative and kinetic analysis of viral shedding and immunological responses in MERS patients representing a broad spectrum of disease severity" | Hospital based | 14 | Viral load was associated with mortality | For | Low | |
| Hong et al (2018). "Predictors of mortality in Middle East respiratory syndrome (MERS)" | Hospital based | 30 | Viral load was significantly associated with mortality | For | Moderate | |
| Corman et al (2016). "Viral Shedding and Antibody Response in 37 Patients With Middle East Respiratory Syndrome Coronavirus Infection" | Hospital based | 37 | Viral load was significantly associated with mortality | For | Moderate | |
| Douglas et al (2018). "Adaptive evolution influences the infectious dose of MERS-CoV necessary to achieve severe respiratory disease" | Animal model (mice) | Not stated | A larger inoculant dose resulted in higher mortality | For | Low | **iii) Infection with a larger dose results in more severe disease** |
| Tao et al (2015). "Characterization and Demonstration of the Value of a Lethal Mouse Model of Middle East Respiratory Syndrome Coronavirus Infection and Disease" | Animal model (mice) | 52 | A larger inoculant dose resulted in higher mortality | For | Low | |

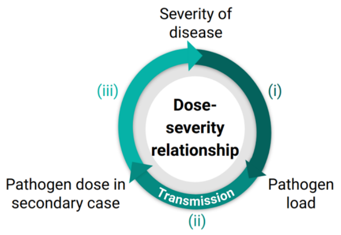

#### Route-severity relationship

| Paper | Study Type | Study size | Key findings | For/against | Strength of evidence | |
|---|---|---|---|---|---|---|
| Widagdo et al (2016). "Differential Expression of the Middle East Respiratory Syndrome Coronavirus Receptor in the Upper Respiratory Tracts of Humans and Dromedary Camels" | Physiological | n/a | Limited replication is possible in the URT due to a lack of DPP4 expression - the receptor used by the virus. Note that they also investigated camels who can be infected in the URT due to having DPP4 expression in those cells | Against | Moderate | **Infection is possible in both the LRT and URT** |
| Raj et al (2013). "Dipeptidyl peptidase 4 is a functional receptor for the emerging human coronavirus-EMC" | Physiological | n/a | DPP4 is thought to be the primary receptor of MERS-CoV infection and DPP4 expression was found only in the bronchiolar epithelial cells and lung tissue. | Against | Moderate | |
| Chan et al (2013). "Tropism of and Innate Immune Responses to the Novel Human Betacoronavirus Lineage C Virus in Human Ex Vivo Respiratory Organ Cultures" | Physiological | n/a | MERS-CoV readily infects LRT tissues. No similar studies have been performed in URT tissues | Against | Low | |
| Assiri et al (2013). "Epidemiological, demographic, and clinical characteristics of 47 cases of Middle East respiratory syndrome coronavirus disease from Saudi Arabia: a descriptive study" | Community based | 47 | The majority of symptoms detected were either systemic or LRT related, not URT | Against | Low | |
| World Health Organisation (2018). "Laboratory testing for Middle East respiratory syndrome coronavirus - revised" | Guidance | n/a | Recommends LRT samples for MERS-CoV PCR testing despite it being more difficult to do | Against | Low | |
| Mackay & Arden (2015). "MERS coronavirus: diagnostics, epidemiology and transmission" | Review | n/a | Studies have found replication in both URT and LRT | For | Low | |
| Vegara-Alert et al (2017). "Searching for animal models and potential target species for emerging pathogens: Experience gained from Middle East respiratory syndrome (MERS) coronavirus" | Review | n/a | MERS has been found to target URT cells in many animal model studies | For | Low | |
| Feikin et al (2015). "Association of Higher MERS-CoV Virus Load with Severe Disease and Death, Saudi Arabia, 2014" | Hospital based | 102 | Significant association between URT viral load and severity | For | Low | |

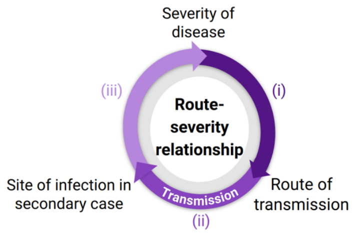

### S8 *Mycobacterium tuberculosis*

| Dose-severity relationship | | | | | | |
|---|---|---|---|---|---|---|
| **Paper** | **Study Type** | **Study size** | **Key findings** | **For/against** | **Strength of evidence** | |
| Shimazaki et al (2018). "Bacterial co-infection and early mortality among pulmonary tuberculosis patients in Manila, The Philippines" | Hospital based | 228 | Bacterial load significantly correlated with two-week mortality | For | Moderate | **i) More severe cases have a higher pathogen load** |
| Du Bruyn et al (2021). "Mycobacterium tuberculosis-specific CD4 T cells expressing CD153 inversely associate with bacterial load and disease severity in human tuberculosis" | Hospital based | 137 | Bacterial load significantly correlated with severity score | For | Moderate | |
| Murthy et al (2018). "Pretreatment chest x-ray severity and its relation to bacterial burden in smear positive pulmonary tuberculosis" | Hospital based | 1354 | Time to positivity (TTP) significantly and negatively correlated with the presence of lung cavities. TPP significantly associated with the extent of disease, although the association was small (a 10-fold increase in TTP was required for an 11.4% decrease in the area affected) | For | Moderate | |
| te Riele et al (2019). "Relationship between chest radiographic characteristics, sputum bacterial load, and treatment outcomes in patients with extensively drug-resistant tuberculosis" | Hospital based | 97 | TTP significantly and negatively correlated with cavity score and negatively associated with disease extent (p=0.071) | For | Moderate | |
| Perrin et al (2010). "Radiological cavitation, sputum mycobacterial load and treatment response in pulmonary tuberculosis" | Hospital based | 95 | TTP significantly and negatively correlated with the presence of lung cavities | For | Moderate | |
| Jones-lopez et al (2016). "Cough Aerosols of Mycobacterium tuberculosis in the Prediction of Incident Tuberculosis Disease in Household Contacts" | Hospital based | 85 | Bacterial load was significantly correlated with a number of severity markers, including length of illness (p=0.04) and sputum volume (p=0.01), but not correlated with the presence of cavities (p=0.65). | For | Moderate | |
| Theron et al (2020). "Bacterial and host determinants of cough aerosol culture positivity in patients with drug-resistant versus drug-susceptible tuberculosis" | Hospital based | 500 | Bacterial load significantly correlated with the presence of lung cavities. | For | Moderate | |
| Palanisamy et al (2008). "Disseminated disease severity as a measure of virulence of Mycobacterium tuberculosis in the guinea pig model" | Animal model (Guinea pigs) | Not stated | Whilst bacterial load was correlated with mortality, the extent to which Mtb had spread outside of the lungs to other vital organs was a more important measure for predicting mortality | For | Low | |
| Acuña-Villaorduña et al (2019). "Host Determinants of Infectiousness in Smear-Positive Patients With Pulmonary Tuberculosis" | Hospital and household based | 245 | Large bacterial load was significantly associated with aerosol positivity | For | Moderate | **ii) A higher pathogen load results in infecting others with a larger dose** |
| Jones-lopez et al (2016). "Cough Aerosols of Mycobacterium tuberculosis in the Prediction of Incident Tuberculosis Disease in Household Contacts" | Hospital based | 85 | A large bacterial load was also significantly associated with median aerosol colony forming unit (CFU) count (p=0.04). Aerosol CFU count was correlated with strength of cough (p=0.008) but was not significantly associated with any measure of TB disease progression (extent of disease(p=0.34), cavitation (p=0.29), sputum volume (p=0.12)). | For | Low | |

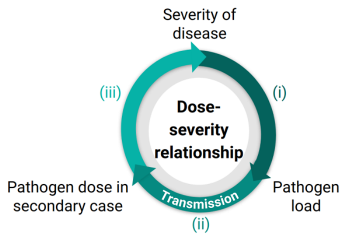

| | | | | | | |
|---|---|---|---|---|---|---|
| Theron et al (2020). "Bacterial and host determinants of cough aerosol culture positivity in patients with drug-resistant versus drug-susceptible tuberculosis" | Hospital based | 500 | Bacterial load was significantly correlated with aerosol CFU count. However, they found a significant positive correlation between strength of cough and aerosol positivity but a negative correlation between symptom score and aerosol positivity (defined by the detection of any aerosol CFUs, p=0.054) | Mixed | | larger dose |
| Fennely & Jones-López (2015). "Quantity and Quality of Inhaled Dose Predicts Immunopathology | Review | n/a | Those infected with a lower inoculant dose are more likely to develop latent TB than if they had been infected with a higher dose | For | Low | **iii) Infection with a larger dose results in more severe disease** |
| Jones-lopez et al (2016). "Cough Aerosols of Mycobacterium tuberculosis in the Prediction of Incident Tuberculosis Disease in Household Contacts" | Hospital based | 85 | Those in contact with a high-aerosol case (an individual with a high aerosol CFU count) were significantly more likely to be infected and to develop TB disease than if they were in contact with a low-aerosol case | For | Low | |

#### Route-severity relationship

| Paper | Study Type | Study size | Key findings | For/against | Strength of evidence | |
|---|---|---|---|---|---|---|
| Fennelly et al (2012). "Variability of Infectious Aerosols Produced during Coughing by Patients with Pulmonary Tuberculosis" | Hospital based | 101 | 96% of culturable Mtb was in particles smaller than 4.7 mu m | Against | Moderate | **Infection is possible in both the LRT and URT** |
| Rohwedder (1994). "Upper respiratory tract tuberculosis. Sixteen cases in a general hospital" | Hospital based | 843 | The URT was involved in less than 2% of TB cases | Against | Moderate | |
| Rohwedder (1994). "Upper Respiratory Tract Tuberculosis" *in* Schlossberg et al (1994) "Tuberculosis" | Review | n/a | URT involvement is likely as a complication of LRT infection, as opposed to infection being initiated in the URT | Against | Low | |
| Tse et al (2003). "Tuberculosis of the Nasopharynx: A Rare Entity Revisited" | Hospital based | 17 | URT involvement is likely as a complication of LRT infection, as opposed to infection being initiated in the URT | Against | Low | |
| Fenneły & Jones-López (2015). "Quantity and Quality of Inhaled Dose Predicts Immunopathology in Tuberculosis" | Review | n/a | Primary URT TB can occur, as shown by multiple case reports, although it is rare | For | Low | |
| Jindal et al (2017). "Upper Respiratory Tract Tuberculosis" *in* Schlossberg et al (2017). "Tuberculosis and Nontuberculous Mycobacterial Infections" | Review | n/a | Found an increase in URT TB reports over the last two decades. | For | Low | |
| King et al (2003). "MR Imaging Features of Nasopharyngeal Tuberculosis: Report of Three Cases and Literature Review" | Hospital based | 3 | Reports three cases of URT TB, where LRT infection was not present | For | Low | |

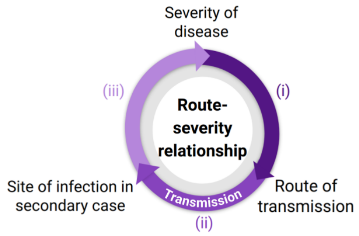

### S9 Rhinovirus

| Dose-severity relationship | | | | | |
|---|---|---|---|---|---|
| **Paper** | **Study Type** | **Study size** | **Key findings** | **For/against** | **Strength of evidence** |
| Ambrosioni et al (2015). "Role of rhinovirus load in the upper respiratory tract and severity of symptoms in lung transplant recipients" | Hospital based | 116 | Viral load tended to be higher in symptomatic patients than asymptomatic patients (p=0.058). High viral load was also associated with symptom score and was significantly associated with sore throat (p<0.001), fever (p=0.011) and cough (p=0.033) | For | Moderate |
| D'Alessio et al (1976). "Transmission of Experimental Rhinovirus Colds in Volunteer Married Couples" | Human challenge | 24 | High viral load was significantly associated with self-rated symptom severity (p=0.0021) | For | Moderate |
| Ng et al (2018). "Viral Load and Sequence Analysis Reveal the Symptom Severity, Diversity, and Transmission Clusters of Rhinovirus Infections" | Hospital based | 976 | High viral load was significantly associated with severity score (p=0.017) | For | Moderate |
| Clark et al (2016). "Viral load is strongly associated with length of stay in adults hospitalised with viral acute respiratory illness" | Hospital based | 149 | Low Ct value significantly associated with duration of hospitalisation (p=0.004) | For | Moderate |
| Message et al (2008). "Rhinovirus-induced lower respiratory illness is increased in asthma and related to virus load and Th1/2 cytokine and IL-10 production" | Hospital based | 25 | High viral load was significantly associated with LRT symptoms, but only in asthmatic patients (p=0.01) | For | Low |
| Takeyama et al (2012). "Rhinovirus load and disease severity in children with lower respiratory tract infections" | Hospital based | 43 | Viral load on admission was not significantly associated with disease severity score when all patients were considered (r=0.126; p=0.421). However, there was a significant association in children >= 11 months of age (r=0.407; p=0.032) | For | Low |
| Piralla et al (2009). "Clinical severity and molecular typing of human rhinovirus C strains during a fall outbreak affecting hospitalized patients" | Hospital based | 250 | For those with HRV-C infection, those with LRT infection had a higher viral load than those with URT infection (p=0.08) and high viral load was associated with pnuemonia (p=0.12) and respiratory distress (p=0.12) | For | Low |
| Sanchez-Codez et al (2021). "Viral Loads and Disease Severity in Children with Rhinovirus-Associated Illnesses" | Hospital based | 2473 | Low viral load significantly decreased the risk of ICU admission (OR 0.97; 95% CI 0.95-0.99) but not the risk of hospitalisation (OR 0.99; 95% CI 0.97-1.01) or need for supplemental oxygen (OR 0.97; 95% CI 0.97-1.00). Note that they don't account for the duration of illness at the time of sample collection | Mixed | |
| Do et al (2016). "Respiratory Syncytial Virus and Other Viral Infections among Children under Two Years Old in Southern Vietnam 2009-2010: Clinical Characteristics and Disease Severity" | Hosptial based | 206 | High viral load was significantly negatively associated with severity in coinfections (r=-0.08,p=0.04), but not in single infections | Against | Low |
| Franz et al (2010). "Correlation of viral load of respiratory pathogens and co-infections with disease severity in children hospitalized for lower respiratory tract infection" | Hospital based | 87 | Viral load not significantly correlated with measures of disease severity | Against | Low |
| Esposito et al (2014). "Impact of rhinovirus nasopharyngeal viral load and viremia on severity of respiratory infections in children" | Hospital based | 50 | High viral load was not significantly associated with hospitalisation (p=0.62), hospital length of stay (p=0.44) or ICU admission (p=0.71) | Against | Low |
| Granados et al (2017). "Influenza and rhinovirus viral load and disease severity in upper respiratory tract infections" | Hospital based | 213 | High viral load was not associated with hospitalisation (p=0.54) or ICU admission | Against | Low |

**i) More severe cases have a higher pathogen load**

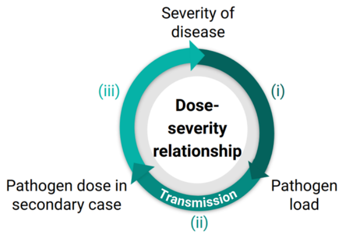

| | | | | | |
|---|---|---|---|---|---|
| Jartti et al (2015). "Rhinovirus-induced bronchiolitis: Lack of association between virus genomic load and short-term outcomes" | Hospital based | 694 | Low Ct was not significantly associated with hospital length of stay (p=0.85) or ICU admission (p=0.4) | Against | Low |
| Houben et al (2010). "Disease severity and viral load are correlated in infants with primary respiratory syncytial virus infection in the community" | Hospital based | 65 | No correlation was found between Ct value and severity score | Against | Low |
| van Elden et al (2007). "Enhanced severity of virus associated lower respiratory tract disease in asthma patients may not be associated with delayed viral clearance and increased viral load in the upper respiratory tract" | Hospital based | 88 | URT viral load was not associated with the severity of LRT symptoms | Against | Low |

| Route-severity relationship | | | | | |
|---|---|---|---|---|---|
| **Paper** | **Study Type** | **Study size** | **Key findings** | **For/against** | **Strength of evidence** |

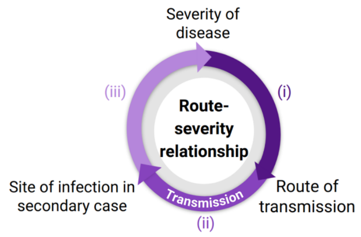

### S10 Respiratory syncytial virus

| Dose-severity relationship | | | | | | |
|---|---|---|---|---|---|---|
| **Paper** | **Study Type** | **Study size** | **Key findings** | **For/against** | **Strength of evidence** | |
| Hijano et al (2019). "Clinical correlation of influenza and respiratory syncytial virus load measured by digital PCR" | Hospital based | 19 | Viral load significantly higher in symptomatic patients and significantly associated with number of symptoms, cough and nasal conjestion but not LRT infection (OR 1.09; 95% CI 0.61-1.95), ICU admission (OR 1.96; 95% CI 0.89-3.94) or death (OR 1.86; 95% CI 0.44-7.79) | For | Moderate | |
| DeVincenzo et al (2010). "Viral load drives disease in humans experimentally infected with respiratory syncytial virus" | Human challenge | 35 | Viral load significantly correlated with disease severity markers: symptom score, physical examination score, and mucus weight | For | Moderate | |
| Fodha et al (2007). "Respiratory syncytial virus infections in hospitalized infants: Association between viral load, virus subgroup, and disease severity" | Hospital based | 81 | Viral load significantly associated with severity (p=0.024) | For | Moderate | |
| Zhou et al (2015). "The impact of viral dynamics on the clinical severity of infants with respiratory syncytial virus bronchiolitis" | Hospital based | 40 | Viral load on day three a signficant risk factor for disease severity but not on day one (OR 1.611; 95% CI 0.62-4.186). Although, viral load on day one was significantly higher in the severe group compared to mild (p=0.034) but not when compared to moderate (data not shown) | For | Moderate | |
| Houben et al (2010). "Disease severity and viral load are correlated in infants with primary respiratory syncytial virus infection in the community" | Hospital based | 82 | Ct value significantly negatively correlated with severity score | For | Moderate | |
| Scagnolari et al (2012). "Evaluation of viral load in infants hospitalized with bronchiolitis caused by respiratory syncytial virus" | Hospital based | 132 | Viral load signficantly correlated with severity score and length of hospital stay | For | Moderate | |
| DeVincenzo et al (2005). "Respiratory Syncytial Virus Load Predicts Disease Severity in Previously Healthy Infants" | Hospital based | 141 | Viral load significantly and independently associated with hospitalisation, respiratory failure and ICU admission | For | Moderate | |
| Fuller et al (2013). "Association of the CT values of real-time PCR of viral upper respiratory tract infection with clinical severity, Kenya" | Hospital based | 523 | Ct value significantly higher in outpatients | For | Moderate | |
| El Saleeby et al (2011). "Respiratory Syncytial Virus Load, Viral Dynamics, and Disease Severity in Previously Healthy Naturally Infected Children" | Hospital based | 219 | Initial viral load significantly associated with duration of hospitalisation. Viral load correlated with ICU admission but only significant when measured on days 2 and 3 (day 1: p=0.55, day 2: p=0.046, day 3: 0.009). | For | Moderate | **i) More severe cases have a higher pathogen load** |
| Walsh et al (2015). "Viral Shedding and Immune Responses to Respiratory Syncytial Virus Infection in Older Adults" | Hospital based | 111 | Viral load significantly correlated with ICU admission but not hospitalisation (URT viral load, p=0.09 and LRT viral load, p=0.06) | For | Moderate | |
| Duncan et al (2009). "Risk Factors for Respiratory Failure Associated with Respiratory Syncytial Virus Infection in Adults" | Hospital based | 58 | Viral load significantly associated with respiratory failure but was not associated with hospitalisation (OR 1.0; 95% CI 0.5-1.7) | Mixed | | |
| Franz et al (2010). "Correlation of viral load of respiratory pathogens and co-infections with disease severity in children hospitalized for lower respiratory tract infection" | Hospital based | 131 | No significant correlation between viral load and any measures of disease severity (e.g. duration of hospital stay, requirement of supplemental oxygen or fever, data not shown) | Against | Low | |

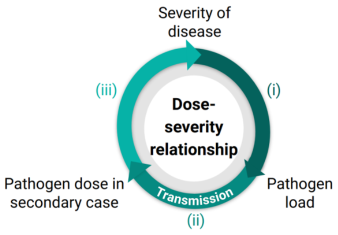

| | | | | | | |
|---|---|---|---|---|---|---|
| Legg et al (2003). "Type 1 and Type 2 Cytokine Imbalance in Acute Respiratory Syncytial Virus Bronchiolitis" | Hospital based | 28 | Viral load not significantly associated with LRT infection (p=0.308, OR not stated) | Against | Low | |
| Mazur et al (2017). "Severity of Respiratory Syncytial Virus Lower Respiratory Tract Infection With Viral Coinfection in HIV-Uninfected Children" | Hospital based | 1330 | No correlation between viral load and life-threatening disease (OR 1.0; 95% CI, 0.94–1.1) or increased length of hospitalisation (OR 1.0; 95% CI, 0.98–1.0). | Against | Low | |
| Garcia-Mauriño et al (2019). "Viral Load Dynamics and Clinical Disease Severity in Infants With Respiratory Syncytial Virus Infection" | Hospital based | 150 | Viral load significantly higher in outpatients compared to inpatients and significantly negatively correlated with symptom score | Against | Moderate | |
| Piedra et al (2017). "The interdependencies of viral load, the innate immune response, and clinical outcome in children presenting to the emergency department with respiratory syncytial virus-associated bronchiolitis" | Hospital based | 79 | Initial viral load significantly correlated with immune response biomarkers and significantly negatively correlated with disease severity | Against | Moderate | |
| Hall and Douglas (1981). "Modes of transmission of respiratory syncytial virus" | Human challenge | 31 | Suggested that less severe disease from fomite transmission compared to large droplet transmission was as a result of a smaller inoculant dose | For | Low | iii) Infection with a larger dose results in more severe disease |
| Hall et al (1981). "Infectivity of respiratory syncytial virus by various routes of inoculation" | Human challenge | 32 | Infection with a larger inoculant dose resulted in an increased viral load. | For | Low | |
| El Saleeby et al (2011). "Respiratory Syncytial Virus Load, Viral Dynamics, and Disease Severity in Previously Healthy Naturally Infected Children" | Hospital based | 219 | No correlation between the number of children in a household (a measure of exposure intensity) and viral load | Against | Low | |
| DeVincenzo et al (2010). "Viral load drives disease in humans experimentally infected with respiratory syncytial virus" | Human challenge | 35 | No correlation between inoculant dose and infection or disease (data not shown) | Against | Low | |
| DeVincenzo et al (2005). "Respiratory Syncytial Virus Load Predicts Disease Severity in Previously Healthy Infants" | Hospital based | 141 | No correlation between number of children in a household or attendance at day care (measures of exposure intensity) and viral load | Against | Low | |

| Route-severity relationship | | | | | | |
|---|---|---|---|---|---|---|
| **Paper** | **Study Type** | **Study size** | **Key findings** | **For/against** | **Strength of evidence** | |
| Johnson et al (2007). "The histopathology of fatal untreated human respiratory syncytial virus infection" | Physiological | 12 | Finds evidence of direct infection in the alveoli during autopsies | For | Low | **Infection is possible in both the LRT and URT** |
| Miles et al (2023). "TLR7 promotes chronic airway disease in RSV-infected mice" | Animal model study (mice) | Not stated | Infection occurs first in the URT before spreading to the LRT | Against | Low | |
| Kulkarni et al (2016). "Evidence of Respiratory Syncytial Virus Spread by Aerosol. Time to Revisit Infection Control Strategies?" | Hospital based | 24 | A "substantial amount" of RSV RNA detected in air filters was in particles <4.7 mu m in diameter | For | Low | **Transmission can occur** |
| Hall and Douglas (1981). "Modes of transmission of respiratory syncytial virus" | Human challenge | 31 | Found transmission through contact transmission but no evidence of long-range aerosol transmission | Against | Low | |

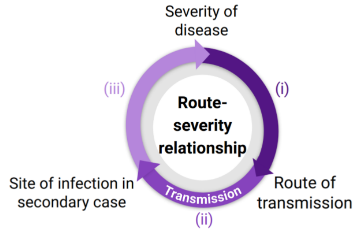

| | | | | | | |
|---|---|---|---|---|---|---|
| Lindsley et al (2010). "Distribution of Airborne Influenza Virus and Respiratory Syncytial Virus in an Urgent Care Medical Clinic" | Hospital based | 285 air samplers | 9% of RSV RNA detected in air filters was in particles less than 4.1 mu m in diameter compared to 42% for influenza | Against | Low | **can occur through aerosols** |
| Grayson et al (2017). "Detection of Airborne Respiratory Syncytial Virus in a Pediatric Acute Care Clinic" | Hospital based | 554 air samplers | Less than 10% of RSV RNA detected in air filters was in particles less than 4.1 mu m in diameter | Against | Low | |

#### S11 SARS-CoV-1

**Dose-severity relationship**

| Paper | Study Type | Study size | Key findings | For/against | Strength of evidence | |
|---|---|---|---|---|---|---|
| Chen et al (2006). "Nasopharyngeal Shedding of Severe Acute Respiratory Syndrome--Associated Coronavirus Is Associated with Genetic Polymorphisms" | Hospital based | 94 | URT viral load significantly correlated with mortality rate (p<0.0001) | For | Moderate | **More severe cases have a higher pathogen load** |
| Cheng et al (2004). "Viral Replication in the Nasopharynx Is Associated with Diarrhea in Patients with Severe Acute Respiratory Syndrome" | Hospital based | 142 | URT viral load on day 10 after symptom onset significantly correlated with mortality (6.2 vs. 1.7 log10 copies/mL; p<0.01). | For | Moderate | |
| Chu et al (2004). "Initial viral load and the outcomes of SARS" | Hospital based | 32 | Initial URT viral load significantly associated with mortality (aOR 1.21 per log10 increase in copies/mL; 95% CI 1.06-1.39) and acute respiratory distress syndrome (aOR 1.19 per log10 increase in copies/mL; 95% CI 1.03-1.38) | For | Moderate | |
| Ng et al (2003). "Quantitative Analysis and Prognostic Implication of SARS Coronavirus RNA in the Plasma and Serum of Patients with Severe Acute Respiratory Syndrome" | Hospital based | 23 | Serum viral load on admission was significantly associated with ICU admission (p<0.005) | For | Moderate | |
| Hung et al (2004). "Viral Loads in Clinical Specimens and SARS Manifestations" | Hospital based | 142 | URT and serum viral load from day 10 to day 15 after symptom onset significantly correlated with diarrhea, oxygen desaturation, mechanical ventilation, hepatic dysfunction and mortality. Note that for patients without these severity markers, the median viral load was 0 log10 copies/mL although they were often still positive by PCR test | For | Low | |
| Wilder-Smith et al (2005). "Asymptomatic SARS Coronavirus Infection among Healthcare Workers, Singapore" | Hospital based | 45 | Wearing a mask was significantly associated with asymptomatic infection (3/6 of asymptomatic patients had worn a mask vs 3/37 of patients with pneumonic SARS). However, other measures of intensity of exposure such as being <3ft from a SARS patient or the length of contact time were not significantly associated with pnuemonia | For | Low | **Infection with a larger dose results in more severe disease** |
| Chu et al (2005). "Viral Load Distribution in SARS Outbreak" | Community based | 321 | Individuals who lived closer to the index case (i.e. in the same residential block) had a higher viral load and increased mortality than those who lived in other blocks. | For | Low | |

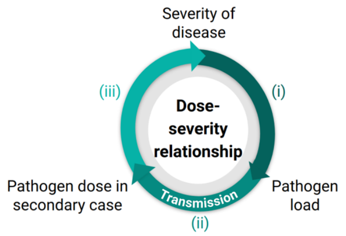

**Route-severity relationship**

| Paper | Study Type | Study size | Key findings | For/against | Strength of evidence |
|---|---|---|---|---|---|

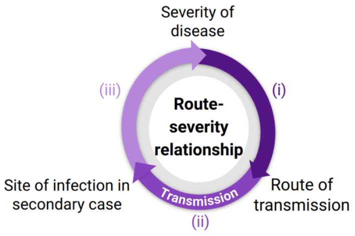

#### S12 SARS-CoV-2

| Dose-severity relationship | | | | | | |
|---|---|---|---|---|---|---|
| **Paper** | **Study Type** | **Study size** | **Key findings** | **For/against** | **Strength of evidence** | |
| Shenoy (2021). "SARS-CoV-2 (COVID-19), viral load and clinical outcomes; lessons learned one year into the pandemic: A systematic review" | Review | n/a | Viral load significantly correlated with mortality and severity of symptoms | For | Moderate | **i) More severe cases have a higher pathogen load** |
| Rabaan et al (2021). "Viral Dynamics and Real-Time RT-PCR Ct Values Correlation with Disease Severity in COVID-19" | Review | n/a | Viral load significantly correlated with mortality and severity of symptoms | For | Moderate | |
| Rao et al (2020). "A Systematic Review of the Clinical Utility of Cycle Threshold Values in the Context of COVID-19" | Review | n/a | Viral load significantly correlated with severity of symptoms | For | Moderate | |
| Chen et al (2021). "SARS-CoV-2 shedding dynamics across the respiratory tract, sex, and disease severity for adult and pediatric COVID-19" | Review | n/a | Viral load significantly correlated with severity of symptoms | For | Moderate | |
| Byrne et al (2020). "Inferred duration of infectious period of SARS-CoV-2: rapid scoping review and analysis of available evidence for asymptomatic and symptomatic COVID-19 cases" | Review | n/a | Viral load significantly higher in those with severe symptoms compared to mild, but similar viral loads between symptomatic and asymptomatic | Mixed | | |
| Walsh et al (2020). "SARS-CoV-2 detection, viral load and infectivity over the course of an infection" | Review | n/a | Viral load significantly higher in those with severe symptoms compared to mild, but similar viral loads between symptomatic and asymptomatic | Mixed | | |
| Fontana et al (2021). "Understanding viral shedding of severe acute respiratory coronavirus virus 2 (SARS-CoV-2): Review of current literature" | Review | n/a | Viral load significantly higher in those with severe symptoms compared to mild, but similar viral loads between symptomatic and asymptomatic | Mixed | | |
| Cevik et al (2021). "SARS-CoV-2, SARS-CoV, and MERS-CoV viral load dynamics, duration of viral shedding, and infectiousness: a systematic review and meta-analysis" | Review | n/a | Viral load similar in asymptomatics and symptomatics | Against | Low | |
| Puchinger et al (2022). "The interplay of viral loads, clinical presentation, and serological responses in SARS-CoV-2 – Results from a prospective cohort of outpatient COVID-19 cases" | Community based | 51 | Moderate symptomatic cases had a significantly higher viral load than both asymptomatic and mildly symptomatic individuals | For | Moderate | |
| Caplan et al (2021). "Clinical characteristics and viral load dynamics of COVID-19 in a mildly or moderately symptomatic outpatient sample" | Community based | 25 | Viral load was higher in moderate vs mild cases, but the difference was not significant (p=0.24) | For | Low | |
| Marks et al (2021). "Transmission of COVID-19 in 282 clusters in Catalonia, Spain: a cohort study" | Household based | 314 | Contacts with a high initial viral load were significantly more likely to develop symptomatic disease (OR per log10 increase in viral load 1·12, 95% CI 1·05–1·20; p=0·0006). However, they found no association between index case viral load and secondary case viral load. | For | Moderate | |

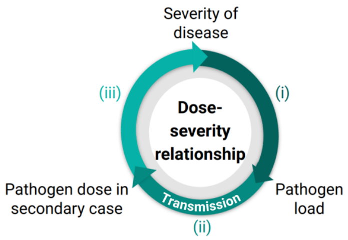

| | | | | | | |
|---|---|---|---|---|---|---|
| Johnson et al (2022). "Viral load of SARS-CoV-2 in droplets and bioaerosols directly captured during breathing, speaking and coughing" | Community based | 17 | Exhaled breath condensate viral load was positively correlated with URT viral load (r=0.5) | For | Moderate | |
| Malik et al (2021). "SARS-CoV-2: Viral Loads of Exhaled Breath and Oronasopharyngeal Specimens in Hospitalized Patients with COVID-19" | Hospital based | 15 | Exhaled breath condensate viral load was not correlated with URT viral load (correlation coefficient R^2 < 0.01) | Against | Low | **ii) A higher pathogen load results in infecting others with a larger dose** |
| Sawano et al (2021). "RT-PCR diagnosis of COVID-19 from exhaled breath condensate: a clinical study" | Hospital based | 50 | EBC viral load was signficantly associated with the requirement of mechanical ventilation and non-significantly associated with the need for oxygen administration (p=0.12) and shortness of breath (p=0.06) | For | Moderate | |
| Sawano et al (2022). "SARS-CoV-2 RNA load and detection rate in exhaled breath condensate collected from COVID-19 patients infected with Delta variant" | Hosptial based | 41 | EBC viral load associated with the need for oxygen administration (p=0.18) | For | Low | |
| Zhou et al (2023). "Viral emissions into the air and environment after SARS-CoV-2 human challenge: a phase 1, open label, first-in-human study" | Human challenge | 36 | Viral load in nasal and throat samples significantly correlated with sampling mask viral load (p<0.0001) | For | Moderate | |
| Dabisch et al (2022). "Comparison of Dose–Response Relationships for Two Isolates of SARS-CoV-2 in a Nonhuman Primate Model of Inhalational COVID-19" | Animal model (Monkeys) | 21 | Relationship between inoculant dose and severity | For | Low | |
| Dabisch et al (2021). "Seroconversion and fever are dose-dependent in a nonhuman primate model of inhalational COVID-19" | Animal model (Monkeys) | 16 | Relationship between inoculant dose and severity | For | Low | |
| Imai et al (2020). "Syrian hamsters as a small animal model for SARS-CoV-2 infection and countermeasure development" | Animal model (Hamsters) | Not stated | Relationship between inoculant dose and severity | For | Low | |
| Lee et al (2020). "Oral SARS-CoV-2 Inoculation Establishes Subclinical Respiratory Infection with Virus Shedding in Golden Syrian Hamsters" | Animal model (Hamsters) | 26 | Relationship between inoculant dose and severity | For | Low | |
| Leist et al (2020). "A Mouse-Adapted SARS-CoV-2 Induces Acute Lung Injury and Mortality in Standard Laboratory Mice" | Animal model (Mice) | Not stated | Relationship between inoculant dose and severity | For | Low | |
| Rosenke et al (2020). "Defining the Syrian hamster as a highly susceptible preclinical model for SARS-CoV-2 infection" | Animal model (Hamsters) | Not stated | Although initially symptoms were more severe in animals given a higher inoculant dose, by day 5 the lower dose animals had more severe and widespread pathology | Against | Low | |
| Trunfio et al (2021). "On the SARS-CoV-2 "Variolation Hypothesis": No Association Between Viral Load of Index Cases and COVID-19 Severity of Secondary Cases" | Household based | 132 | No relationship between the viral load of the index and the severity of secondary cases | Against | Low | **iii) Infection [illegible]** |

| | | | | | | |
|---|---|---|---|---|---|---|
| Raoult et al (2020). "Coronavirus infections: Epidemiological, clinical and immunological features and hypotheses" | Review | n/a | A larger inoculant dose can overwhelm the host's defence and causing a state of immunosuppression | For | Low | **iii) Infection with a larger dose results in more severe disease** |
| Kikkert et al (2020). "Innate Immune Evasion by Human Respiratory RNA Viruses" | Review | n/a | The initial immune response may be insufficient to clear a high dose, leading to the use of a second line of defence which triggers increased inflammation | For | Low | |
| Maltezou et al (2020). "SARS-CoV-2 Infection in Healthcare Personnel With High-risk Occupational Exposure: Evaluation of 7-Day Exclusion From Work Policy" | Hospital based | 3,395 | Healthcare workers with high-risk exposure (close contact with a COVID-19 case with neither party wearing a mask) were significantly more likely than those with moderate- or low-risk exposures to develop symptoms (31.9%, 22.6%, and 15.8%, respectively; $p < 0.001$) and to be hospitalised (0.8%, 0.4%, and 0.1%, respectively; $p < 0.001$) | For | Moderate | |
| Zhang et al (2020). "Factors associated with asymptomatic infection in health-care workers with severe acute respiratory syndrome coronavirus 2 infection in Wuhan, China: a multicentre retrospective cohort study" | Hospital based | 424 | Healthcare workers who had performed tracheal intubation or extubation were significantly more likely to have symptomatic infection (OR 4.057; $p=0.026$) and that the consistent use of respirators significantly decreased the risk of symptomatic infection (OR 0.369; $p = 0.001$). | For | Moderate | |
| Bielecki et al (2021). "Social Distancing Alters the Clinical Course of COVID-19 in Young Adults: A Comparative Cohort Study" | Community based | 508 | NPIs resulted in a reduction in the proportion of cases that were symptomatic. In the two barracks where the outbreak occured before NPIs were implemented, 102 individuals had symptomatic infection and 113 had confirmed infection by PCR test. In the barrack where individuals only became infected after NPIs were implemented, no symptomatic cases were detected despite 13 individuals testing positive. | For | Low | |
| Gandhi et al (2020). "Masks Do More Than Protect Others During COVID-19: Reducing the Inoculum of SARS-CoV-2 to Protect the Wearer" | Review | n/a | NPIs resulted in a reduction in the proportion of cases that were symptomatic. Whilst the typical rate of asymptomatic infection was estimated by the CDC in july 2020 to be around 40%, asymptomatic infection rates were estimated to be around 80% when masks were used universally. | For | Low | |
| Soriano et al (2021). "Main differences between the first and second waves of COVID-19 in Madrid, Spain" | Community based | 1569 | NPIs resulted in a reduction in the proportion of cases that were symptomatic. In the first wave (no NPIs), 34/122 (27.8%) of those who tested positive had clinical symptoms. In the second wave (with NPIs), only 5/47 (10.6%) had clinical symptoms) | For | Low | |
| Chan et al (2020). "Surgical Mask Partition Reduces the Risk of Noncontact Transmission in a Golden Syrian Hamster Model for Coronavirus Disease 2019 (COVID-19)" | Animal model (Hamsters) | 27 | They found that the presence of a mask barrier not only reduced transmission but also reduced severity in the animals that were infected | For | Low | |

#### Route-severity relationship

| Paper | Study Type | Study size | Key findings | For/against | Strength of evidence | |
|---|---|---|---|---|---|---|
| Leding et al (2022). "Detection of SARS-CoV-2 in exhaled breath from non-hospitalized COVID-19-infected individuals" | Hospital based | 111 | Asymptomatics can also transmit disease via aerosols but symptomatic individuals were significantly more likely to have SARS-CoV-2 detected in EBC than asymptomatic individuals (OR 4.4; p=0.017) | For | Moderate | **i) More severe disease results in increased aerosol production** |
| Sawano et al (2021). "RT-PCR diagnosis of COVID-19 from exhaled breath condensate: a clinical study" | Hospital based | 50 | The detection of viral RNA in EBC was significantly associated with the need for oxygen administration (p<0.01), the need for mechanical ventilation (p=0.04), cough (p<0.01) and fever (p=0.01). | For | Moderate | |
| Zhou et al (2023). "Viral emissions into the air and environment after SARS-CoV-2 human challenge: a phase 1, open label, first-in-human study" | Human challenge | 36 | Volunteers were inoculated intranasally. VIral RNA was detected in 109 (43%) of 252 mask samples from 17 participants and viable SARS-CoV-2 was collected from breaths captured in 16 of those masks. Individuals who reported the highest symptom scores were not those who emitted the most virus. For example, one participant who was in the high symptom score group emitted relatively little virus, whereas another participant who was asymptomatic emitted large amounts of virus | Against | Moderate | |
| Johnston et al (2021). "Development of a coronavirus disease 2019 nonhuman primate model using airborne exposure" | Animal model (Monkeys) | 11 | Aerosol inoculation results in severe disease more often than intranasal inoculation | For | Low | **iii) Infection in the LRT (i.e. via aerosols) results in more severe disease than URT infection** |
| Port et al (2021). "SARS-CoV-2 disease severity and transmission efficiency is increased for airborne compared to fomite exposure in Syrian hamsters" | Animal model (Hamsters) | 36 | Aerosol inoculation results in severe disease more often than intranasal inoculation | For | Low | |
| Bixler et al (2022). "Exposure Route Influences Disease Severity in the COVID-19 Cynomolgus Macaque Model" | Animal model (Monkeys) | 16 | Aerosol inoculation results in severe disease more often than intranasal inoculation | For | Low | |

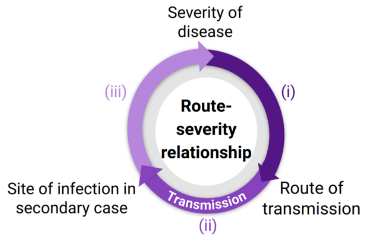

### S13 Variola virus

**Dose-severity relationship**

| Paper | Study Type | Study size | Key findings | For/against | Strength of evidence |
|---|---|---|---|---|---|
| Rao et al (1968). "Epidemiological studies in smallpox. A study of intrafamilial transmission in a series of 254 infected families." | Household based | 254 | Viral shedding correlated with severity | For | Low |
| Smith (2013). "Smallpox: can we still learn from the journey to eradication?" | Review | n/a | Smallpox is thought to have a long latent period because initial infection goes undetected by innate immune system. By the time the virus is detected, mass replication has occured resulting in a high viral load | Against | Low |

**i) More severe cases have a higher pathogen load**

**iii) Infection with a larger dose results in more severe disease**

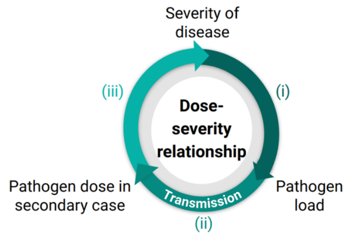

**Route-severity relationship**

| Paper | Study Type | Study size | Key findings | For/against | Strength of evidence |
|---|---|---|---|---|---|
| Hopkins (1983). "Princes and peasants: smallpox in history" | Review | n/a | Suggestion that natural infection is more severe than variolation due to aerosols reaching the lungs, instead of the nose or throat | For | Low |
| Hahon & Wilson (1960). "Pathogenesis of variola in Macaca irus monkeys" | Animal model (Monkeys) | Not stated | Aerosol inoculation resulted in initial infection of the lungs, before spread to the URT | For | Low |
| Katler et al (1979). "Experimental smallpox in chimpanzees" | Animal model (chimpanzee) | 4 | More severe disease after long-range aerosol infection than after contact transmission or direct inoculation | For | Low |
| Sarkar et al (1973). "Virus excretion in smallpox" | Household based | 328 | Only 4/34 of contacts with positive throat swabs developed symptomatic disease | For | Low |

**iii) Infection in the LRT (i.e. via aerosols) results in more severe disease than URT infection**

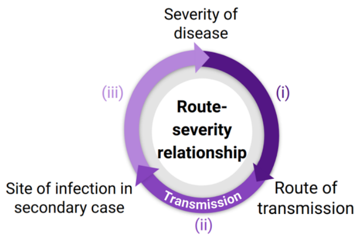

### S14 Variola zoster virus

| Dose-severity relationship | | | | | | |
|---|---|---|---|---|---|---|
| **Paper** | **Study Type** | **Study size** | **Key findings** | **For/against** | **Strength of evidence** | |
| Malavige et al (2008). "Viral Load, Clinical Disease Severity and Cellular Immune Responses in Primary Varicella Zoster Virus Infection in Sri Lanka" | Hospital based | 34 | Serum viral loads were significantly higher in patients with moderate to severe infection compared to thosewith mild infection (p<0.001) | For | Moderate | **i) More severe cases have a higher pathogen load** |
| Poulsen et al (2002) | Household based | 165 | The mean number of pox was signifcantly higher in secondary cases infected within the household than in primary cases (p<0.01), however there was no difference in fever response. | Mixed | | **iii) Infection with a larger dose results in more severe disease** |
| Ross et al (1962). "Modification of Chicken Pox in Family Contacts by Administration of Gamma Globulin" | Household based | Not stated | The mean number of pox was higher in secondary cases infected within the household than in primary cases | For | Low | |

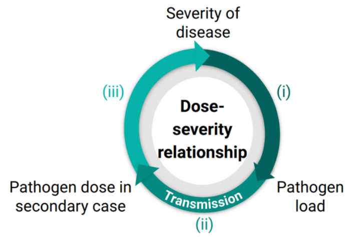

| Route-severity relationship | | | | | |
|---|---|---|---|---|---|
| **Paper** | **Study Type** | **Study size** | **Key findings** | **For/against** | **Strength of evidence** |

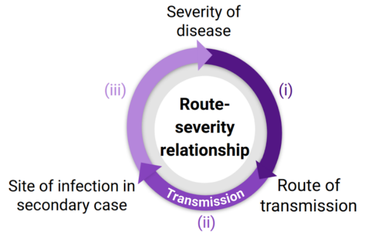

#### S15 *Yersinia pestis*

**Dose-severity relationship**

| Paper | Study Type | Study size | Key findings | For/against | Strength of evidence | |
|---|---|---|---|---|---|---|
| Guinet et al (2015). "Dissociation of Tissue Destruction and Bacterial Expansion during Bubonic Plague" | Animal model (Mice) | Not stated | Bacterial loads in lymph nodes correlated with bubonic plague mortality | For | Low | **i) More severe cases have a higher pathogen load** |
| Druett et al (1956). "Studies on respiratory infection: II. The influence of aerosol particle size on infection of the guinea-pig with Pasteurella pestis" | Animal model (Guinea pig) | Not stated | Animals inoculated with a larger dose did not cause increased mortality in those they infected. The number of deaths increased with the dose, but the number of animals infected at each dose was not stated meaning the mortality rate cannot be compared | Against | Low | **iii) Infection with a larger dose results in more severe disease** |
| Parry (1956). "An interference phenomenon caused by Pasteurella pestis" | Animal model (Rat) | 160 | A larger inoculant dose resulted in decreased mortality and their deaths were delayed in fatal cases | Against | Low | |

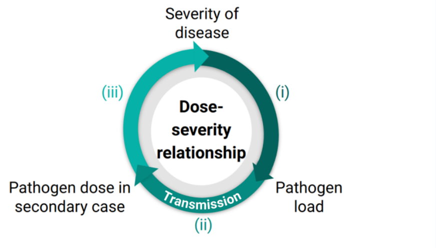

**Route-severity relationship**

| Paper | Study Type | Study size | Key findings | For/against | Strength of evidence | |
|---|---|---|---|---|---|---|
| Petrie (1911). "An epidemiological review of the epidemic of pneumonic plague in northern China, 1910 to 1911" *in* "Report of the International plague conference held at Mukden, April, 1911", pg. 408-468 | Review | n/a | Person-to-person transmission of pnuemonic plague is well-documented to result in pneumonic plague | For | High | **General** |
| Drancourt et al (2006). "Yersinia pestis as a telluric, human ectoparasite-borne organism" | Review | n/a | Person-to-person transmission of pnuemonic plague is well-documented to result in pneumonic plague | For | High | |
| Kool et al (2005). "Risk of Person-to-Person Transmission of Pneumonic Plague" | Review | n/a | Person-to-person transmission of pnuemonic plague is well-documented to result in pneumonic plague | For | High | |
| Dennis et al (1999). "Plague manual: epidemiology, distribution, surveillance and control" | Review | n/a | Person-to-person transmission of pnuemonic plague is well-documented to result in pneumonic plague | For | High | |
| Chandler et al (1997). "Plague" *in* Connor et al (1997). "Pathology of infectious diseases" | Review | n/a | Person-to-person transmission of pnuemonic plague is well-documented to result in pneumonic plague | For | High | |
| Perry & Fetherston (1997). "Yersinia pestis--etiologic agent of plague" | Review | n/a | Person-to-person transmission of pnuemonic plague is well-documented to result in pneumonic plague | For | High | |
| Strong & Teague (1911). "The infectivity of the breath" *in* "Report of the International plague conference held at Mukden, April, 1911", pg.83-87 | Hospital based | Not stated | Y. pestis was detected in the air after patients with pneumonic plague coughed | For | Moderate | **i) More severe disease results in increased aerosol production** |
| Meyer (1961). "Pneumonic plague" | Review | n/a | Aerosol transmission results in pnuemonic plague, whereas infection via large droplets results in URT plague | For | Moderate | **iii) Infection in the LRT (i.e. via aerosols) results in more severe** |
| Agar et al (2009). "Characterization of the rat pneumonic plague model: infection kinetics following aerosolization of Yersinia pestis CO92" | Animal model (Rat) | 50 | Rats infected via the aerosolised route developed pneumonic plague, and were able to transmitt pneumonic plague to uninfected rats | For | Moderate | |

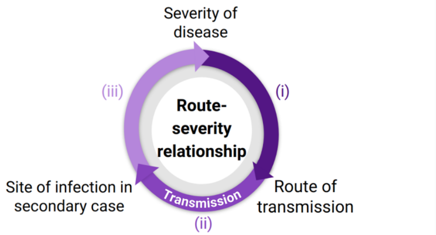

| | | | | | | |
|---|---|---|---|---|---|---|
| Druett et al (1956). "Studies on respiratory infection: II. The influence of aerosol particle size on infection of the guinea-pig with Pasteurella pestis" | Animal model (Guinea pig) | Not stated | Inoculation with large droplets resulted in URT infection, whereas aerosols lead to LRT infection. In addition, they found that mortality was four times higher in secondary cases who were in contact with aerosol-infected animals. | For | Low | disease than URT infection |
